# Patient, carer and clinical stakeholder views on using artificial intelligence and machine learning for the histopathological diagnosis of duodenal biopsies

**DOI:** 10.64898/2026.11.16.26368196

**Authors:** Joel Baby, Florian Jaeckle, Josephine Williams, Keira Shepherd, Heidi Urwin, Elizabeth Soilleux

## Abstract

**Objective:** Biopsies are examined under the microscope by a pathologist, giving a diagnosis, which guides treatment and predicts prognosis. A shortage of pathologists and increasing workload leads to diagnostic delays, while for duodenal (small intestinal) biopsies, variable diagnostic concordance between pathologists limits accuracy. These challenges have motivated us to develop an artificial intelligence (AI)-mediated approach for the automated diagnosis of up to 80% of duodenal biopsies (i.e., most cases of the two most frequent diagnoses: coeliac disease and “normal”). We set out to understand patient, carer and stakeholder views regarding the adoption of such technology.

**Methods and Analysis:** We conducted mixed-methods stakeholder engagement (February 2024 to June 2026). We ran a national online patients’ and carers’ questionnaire (n=231 respondents) and four UK focus-group workshops, one of which additionally captured Likert-scale attitudes^1^ immediately before and after a structured educational session (n=34 matched pairs). Healthcare professionals completed an online questionnaire (n=37) and took part in a structured workshop. Quantitative data were summarised descriptively; free-text and workshop data were analysed thematically and triangulated across all sources.

**Results:** Most patients and carers supported AI-reliant diagnosis (68.4%) and would strongly trust systems with formal regulatory approval (66.7%). Most wanted to be informed if AI was used (73.6%), but only 52.4% felt explicit permission was necessary. Qualitative themes centred on clear and available evidence of accuracy, human oversight, transparent communication and clear terminology, with the term ‘machine learning’ being preferred to ‘artificial intelligence’. Agreement (‘agree’ or ‘strongly agree’ combined) with statements relating to trust, acceptance and perceived benefit of AI was significantly higher after a structured educational session (38.2% to 67.6%, 85.3% to 97.1%, and 52.9% to 97.1% respectively; all p≤0.003). Healthcare professionals favoured an AI ‘assistant’ model over full automation, citing the need for validation, explainability, data governance and workflow integration. Patients also preferred some level of human oversight.

**Conclusion:** Patients and healthcare professionals are cautiously supportive of AI-assisted diagnosis of duodenal biopsies, conditional upon demonstration of accuracy, appropriate human oversight and transparent communication. Attitudes towards AI-assisted diagnosis were more favourable after structured explanation.

**Key Messages:** *What is already known on this topic:* Very little is known about patients’, carers’ or healthcare professionals’ views about the automation of diagnosis in pathology, using AI-based software solutions.

*What this study adds:*

⇒ This is the first study to investigate patient, carer and healthcare professional attitudes specifically to AI-mediated duodenal biopsy diagnosis. Around two-thirds of respondents supported AI-assisted diagnosis, contingent upon clear and available evidence of accuracy, appropriate human oversight and transparent communication, together with appropriate regulatory approval of the technology.
⇒ Attitudes were markedly more favourable after a single structured educational session: agreement that AI could improve duodenal biopsy diagnosis nearly doubled and trust in healthcare AI rose substantially, suggesting that public views of AI in diagnosis can be influenced by clear explanation.
⇒ Patients and healthcare professionals both favour some ongoing level of human oversight of any AI-based diagnostic approach.

*How this study might affect research, practice or policy:*

⇒ Our study suggests that a phased, AI-assistant-first deployment model with defined validation and audit checkpoints would be most readily acceptable when introducing diagnostic AI into histopathology.
⇒ It indicates that appropriate information and education, for example, a patient and public involvement and engagement (PPIE) session, can increase trust in AI in healthcare.
⇒ It shows that patients and the public are more trusting of technology for which the term machine learning is used, rather than artificial intelligence (AI).
⇒ It offers a stakeholder-engagement template for use by others introducing AI-based diagnostics.

## Introduction

Biopsies are examined under the microscope by a clinically qualified histopathologist (“pathologist”), giving a diagnosis, which guides treatment and predicts prognosis, e.g., for cancer. Delays in Pathology impact clinical services, worsen patient outcomes, and increase patient anxiety^2 3^. Pathology workloads are rising, driven by increasing cancer incidence (with a 20% rise predicted 2024-2039^4–6^) and specimen complexity, as more parameters are needed to guide prognosis and treatment^5 7 8^. Combined with pathologist shortages, this worsens turnaround times, with a 2023 UK study showing wide variation in skin biopsy reporting times (1-28 weeks, with the longest departmental average being 5.8 weeks)^9^, compared with a Royal College of Pathologists (RCPath) target that 80% of cases should be reported within 7–10 calendar days^10^. In 2018, 78% of UK pathology departments had vacant consultant posts, mirroring the international picture^8 11^; in 2016, half of NHS departments used locums (£17 million annually) and 45% subcontracted biopsy diagnosis (£10 million annually)^5^. The number of UK patients waiting more than 6 weeks for a pathology diagnosis rose approximately 17% annually between 2010 and 2016^1^. In North America, at least half of pathologists report burnout, with a third considering retirement and a further third a career change^12 13^.

Various industries have embraced artificial intelligence (AI)-mediated image recognition, but uptake in healthcare has been slower. Unlike histopathology, other clinical laboratory disciplines (haematology, biochemistry, immunology) have automated much of their diagnosis^14 15^. Radiology provides the best parallel to histopathology, with Oxipit (Vilnius, Lithuania) obtaining regulatory approval (CE mark under Regulation (EU) 2017/745) for automated diagnosis of normal chest X-rays, sending all abnormal, and some normal, chest X-rays to a radiologist^16–18^. Funded by Innovate UK, some UK histopathology departments have transitioned from glass slides under microscopes to digitised biopsy images on screens, a shift that could similarly mitigate pathologist shortages, costs and regional variation in diagnostic accuracy, which depends on local histopathologists’ expertise^19–25^.

AI-mediated reading of some digitised biopsy slide images has previously been suggested as a mitigation for histopathology workforce shortages ^6 26 27^. In developing a solution, we recognise that applying such a technology to more straightforward, “low risk” biopsy types (not cancers) is most likely to gain market traction. Duodenal (small bowel) endoscopic biopsies are an ideal starting point:

1. <1% show malignancy ^28^, minimising clinical/medicolegal risk.
2. This biopsy type (>400,000 per annum) consumes >40,000 hours of pathologist time annually^29^, hence automation can have a significant impact.
3. Its relatively consistent and recognisable patterns^30^ should make it well suited to a software solution.
4. A software solution that can make just two diagnoses (coeliac disease (CD) and “no abnormality”) could decrease pathologists’ duodenal biopsy workloads by around 80% ^28^.
5. Current diagnostic accuracy for duodenal biopsies is low (with a 20-25% disagreement rate between histopathologists^19–25 31^, contrasting with 3–6% for biopsies from other body sites^32–34^), but this could be improved with a software solution.

The current lack of diagnostic accuracy for duodenal biopsies potentially leads to systematic underdiagnosis of one particular condition: coeliac disease ^31^. Together with low awareness of CD among both the public and clinicians, this contributes to misclassification and diagnostic delay^35^. Population screening studies estimate that around 1% of UK adults and children have coeliac disease^36 37^, yet only a quarter to a third of these have a formal diagnosis^38 39^. With a UK population of around 69.5 million^40^, this indicates that up to an estimated half a million people with coeliac disease are living without a diagnosis. Untreated CD causes substantial morbidity (abdominal pain, diarrhoea, fatigue, mouth ulcers, skin rashes, “brain fog”, peripheral neuropathy, ataxia) and longer-term complications including cancer, lymphoma, vitamin deficiency, anaemia, osteoporosis and infertility^41–44^. A US Air Force cohort study found a near four-fold increase in mortality over 45 years’ follow-up in those with undiagnosed CD^45^, whereas an Edinburgh study found that CD diagnosed in childhood carried a mortality rate similar to the general population^46^. Diagnosis is key in CD because it is simply and effectively treated with a gluten-free diet. Histopathology is currently regarded as the gold standard for diagnosing CD^35^.

Recent advances in digital pathology and AI, specifically machine learning, could improve diagnostic accuracy, efficiency and capacity, helping mitigate international pathologist shortages^25 28 47–50^. Deep learning applied to histological slides can detect complex morphological patterns and assist with disease classification, even matching pathologist-level performance^49^. We have developed a software system for duodenal biopsy histopathological diagnosis with performance comparable to experienced pathologists^49^ and have made its output interpretable by reporting parameters pathologists themselves assess (e.g. cell counts)^48^. Beyond regulatory approval and compliance with cybersecurity, data protection and consent frameworks, successful clinical adoption requires support, or at least acceptance, from key stakeholder groups. We therefore engaged patients and carers and healthcare professionals through a series of bespoke events. Because CD is a key diagnosis for our system and the most common specific pathology seen in the duodenum, and Coeliac UK is a well-organised patient support organization^51^, most of our PPIE has been conducted with CD patients.

Our research questions were: (1) What benefits, risks and prerequisites do (a) patients and carers and (b) healthcare professionals perceive in AI-enabled duodenal biopsy diagnosis? (2) Where do the views of patient and healthcare professionals perspectives converge or diverge, and how might this inform a feasible, ethical deployment plan? (3) Does structured explanation of the technology and its safeguards change patients and carers attitudes? To our knowledge, this is the first study to capture patient, carer and healthcare professional perspectives on AI-assisted histopathological diagnosis of duodenal biopsies, extending an approach previously applied to prostate cancer histopathology^52^.

## Methods

### Participant Recruitment and Data Collection

We conducted scoping surveys to capture patients and carers’ and healthcare professionals’ perspectives, including perceived benefits, risks and operational considerations, on integrating our AI approach into the diagnostic pathway for duodenal biopsies. Data collection combined methods across seven activities (Table 1): for patients and carers, one national online questionnaire (Supplementary Figure 1) and four in-person focus-group workshops; for healthcare professionals, one online questionnaire and one online workshop. Data were collected and analysed anonymously; participation was voluntary throughout, with implied consent given by completing the survey or attending in person. Healthcare professionals’ workshop participants were invited from a broad range of clinical and scientific backgrounds (histopathology, dietetics, gastroenterology, general practice, oncology and ‘other’). Blank questionnaires/ discussion guides and full quantitative results are provided in Supplementary Methods (Supplementary Figures 1–9) and Supplementary Results (Supplementary Tables 1–12) respectively. As these were stakeholder engagement activities rather than research on identifiable individuals, formal ethical approval was not required.

**Table 1.** Summary of data collection events. Our seven stakeholder engagement activities are listed in date order within each group (patients and carers, events 1–5; healthcare professionals, events 6–7). Participant numbers are those who took part or responded. Ages are means (range) where recorded; “not recorded” means the information was not collected. Abbreviations: AI, artificial intelligence; GP, general practitioner; HCPC, Health and Care Professions Council; IBMS, Institute of Biomedical Science; IfM, Institute for Manufacturing; NHS, National Health Service; RCPath, Royal College of Pathologists.

| **Event no.** | **Activity** | **Date** | **Location** | **Participant number** | **Participants’ mean ages (and range; number not recorded) for patients and carers; or number (percentage) of individuals with each type of role for healthcare professionals** | **Participant genders (male; female; other; number not recorded)** | **Session lead(s)** | **Key topics** | **Supporting materials** |
|---|---|---|---|---|---|---|---|---|---|
| ***Patients and Carers*** |  |  |  |  |  |  |  |  |  |
| 1 | National online questionnaire (patients & carers) | Feb 2024-Dec 2025 | Online, UK (advertised to Coeliac UK members) | 231 responses (from *circa* 60,000 eligible members) | 62.9 (20–91; 5 not recorded) | 52; 177; 2; 0 | Coeliac UK | Awareness, attitudes, acceptance, transparency, permission, trust, demographics | Questionnaire: Supplementary Fig. 1; results: Supplementary Tables 1–2; data in Figure 1 |
| 2 | Focus-group workshop (patients & carers) | March 2024 | Stevenage, UK | n=24 | 55.8 (27–83; 3 not recorded) | Not recorded | Coeliac UK | Diagnostic journeys, perceived AI benefits/risks, preferred oversight, communication preferences | Questionnaire: Supplementary Fig. 2; results: Supplementary Table 4 |
| 3 | Focus-group workshop (patients & carers) | June 2025 | Bury St Edmunds, UK | n=20 | Not recorded (anonymous questionnaire) | Not recorded | Lyzeum Ltd | Diagnostic journeys, perceived AI benefits/risks, preferred oversight, communication preferences. More emphasis on patient advocacy and pathway to clinical adoption | Questionnaire: Supplementary Fig. 3; results: Supplementary Table 5 |
| 4 | Focus-group workshop (patients & carers) | September 2025 | Edinburgh, UK | n=33 (27 responses received and analysed) | 60.7 (26–82; 3 not recorded) | Not recorded | Coeliac UK | Diagnostic journeys, perceived AI benefits/risks, preferred oversight, communication preferences | Questionnaire: Supplementary Fig. 4; results: Supplementary Table 6 |
| 5 | Focus-group workshop and educational session with pre/post questionnaire (patients & carers) | June 2026 | Cambridge, UK | n=34 | 58.7 (23–84; 2 not recorded) | 13; 21; 0; 0 | Coeliac UK | Matched Likert-scale attitudes before and after a structured education session about the envisaged AI-based diagnostic solution and the validation/regulation needed around its introduction; demographics (age, gender, education, family history of CD, personal biopsy history, healthcare/tech occupation) | Questionnaire: Supplementary Figures 5-7; results: Supplementary Tables 7-9 |
| ***Clinical and managerial stakeholders*** |  |  |  |  |  |  |  |  |  |
| 6 | Online workshop (healthcare professionals) | June 2024 | Online, UK | n=7 | Histopathologist 4 (57.1%; one also an NHS manager); gastroenterologist 1 (14.3%); NHS manager 2 (28.6%) | Not recorded | IfM Engage/Lyzeum Ltd | Barriers across 4 domains: trust/explainability, privacy/security, digital integration, usability/deskilling (prioritisation exercise + plenary discussion) | Questionnaire: Supplementary Fig. 9; results: Supplementary Table 12 |
| 7 | Online questionnaire (healthcare professionals) | May–Nov 2025 | Online, UK (advertised via RCPath, IBMS, Coeliac UK, HCPC, The Pathologist) | 37 responses | Histopathologist 9 (24%); dietitian 5 (14%); gastroenterologist 4 (11%); GP 1 (3%); other 10 (27%, including 1 oncologist); not recorded 8 (22%) | Not recorded | Lyzeum Ltd | AI assistance vs. automation preferences, perceived obstacles/benefits, workflow impact | Questionnaire: Supplementary Fig. 8; results: Supplementary Tables 10-11 |

### Patient and Public Involvement and Engagement (PPIE)

Coeliac UK, a national patient charity, contributed to the design and interpretation of PPIE study materials and events, and facilitated recruitment. Two patients on the Lyzeum Ltd Steering Committee contributed to discussions about PPIE event formats. Individual patients and carers, as well as clinical and managerial stakeholders, participated voluntarily in the workshops and questionnaires described in this study.

### Analysis

Quantitative data were summarised descriptively (proportions, means, medians). Free-text responses, focus-group materials and workshop outputs were analysed using inductive thematic analysis^53^. Matched pre/post-workshop responses (n=34) for seven Likert-scale items^1^ were compared using the Wilcoxon signed-rank test; a two-sided *p<*0.05 was considered significant. Findings across all sources were integrated via triangulation to identify convergence, complementarity and divergence between patients and carers and healthcare professionals, and across the four focus-group sites and national survey. Associations between demographics (age, gender, education) and six attitude/trust items (Q1– Q6) in the national survey (n=231) were tested with Kruskal-Wallis (age, education) and chi-square (gender; Monte Carlo permutation p-values where expected counts <5) tests, plus Spearman correlations for the two ordinal items (Q2, Q6); Benjamini–Hochberg correction was applied across all 22 tests (α=0.05). Cohort demographics were compared with 2021 UK Census values via exact binomial tests, using R version 4.6.0.

## Results

### Data collection Approaches

Four UK in-person patient and carer events, in Stevenage, Bury St Edmunds, Edinburgh and Cambridge, one online workshop for healthcare professionals, and two online questionnaires (one for patients and carers; one for healthcare professionals) ran over twenty-nine months (Table 1).

### Coeliac UK national survey for patients and carers

Respondents (n=231) were disproportionately female (76.6% vs. 51.0% of the general population^54^) and more highly educated than the UK general population (57.4% educated to at least degree level or equivalent vs. 33.8% nationally^54 55^), and spanned a wide age range (mean 62.9, median 65, range 20–91; Supplementary Table 1). The preponderance of female respondents is in keeping with our observation that approximately two thirds of the biopsy samples showing coeliac disease in our recent studies come from female patients^49 50^, and also consistent with nationally reported gender ratios in this condition^56^.

Attitudes to AI in daily life were broadly positive (56.7% ‘very positive’/’positive’ combined), 12.1% negative (very negative/negative combined), and 31.2% neutral (Figure 1). 68.4% would accept an AI-reliant diagnosis, 26.0% were unsure and 5.6% were opposed; subsequent workshops clarified this acceptance was conditional on clear and available evidence of accuracy, regulatory approval and availability of human oversight or second opinion. 73.6% felt patients should be told if AI was used; free-text responses emphasised transparency and context (e.g. performance relative to conventional practice). 52.4% felt explicit permission should be sought from patients if AI were to be used in the diagnosis of their biopsy. However, many viewed AI as analogous to other validated laboratory technologies, especially where a clinician remained accountable. When described as approved by “all necessary regulatory bodies,” 66.7% strongly or very strongly trusted the use of AI in diagnosis (Figure 1, Supplementary Table 1).

**Figure 1:**
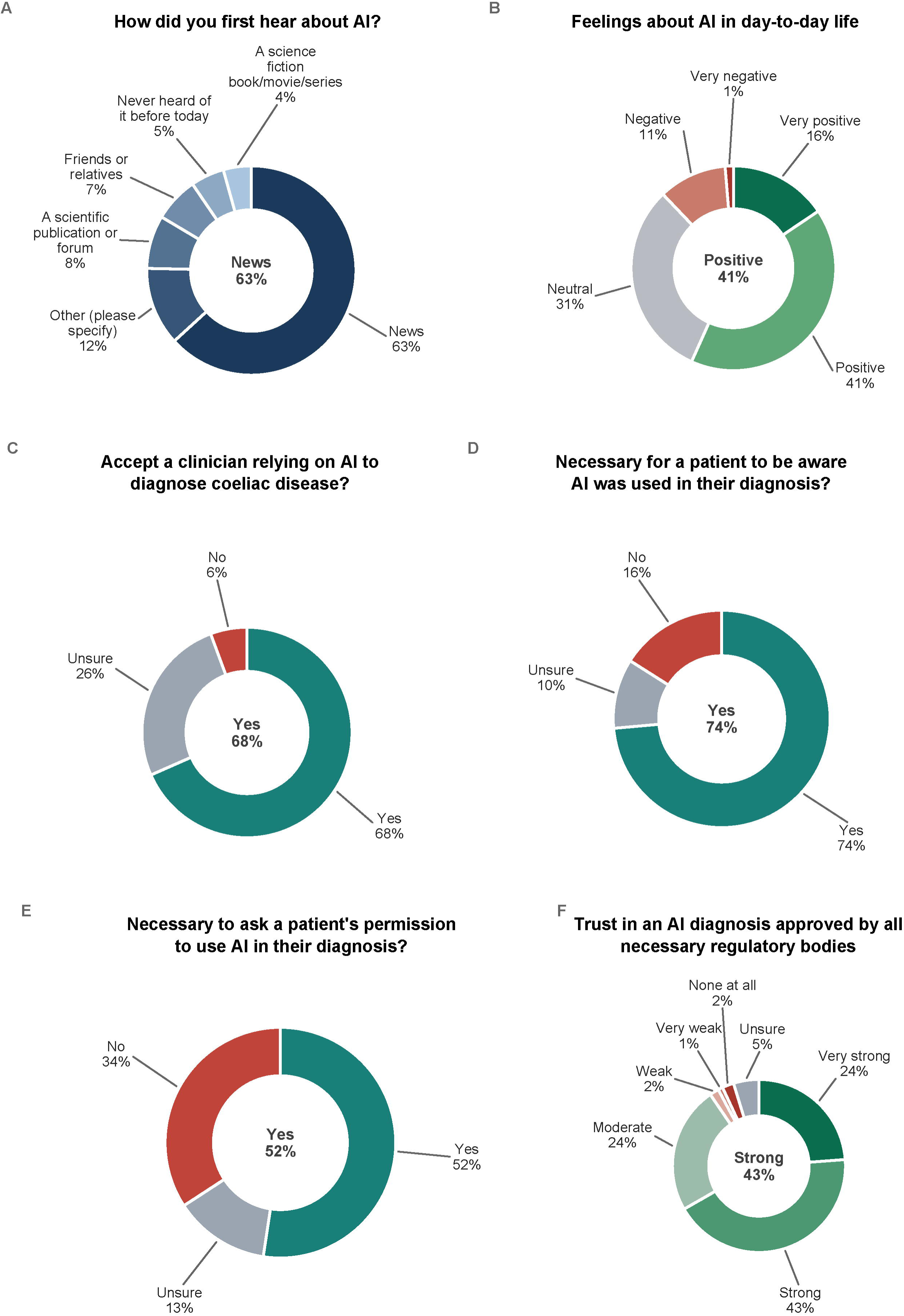
Public perceptions and attitudes toward the integration of artificial intelligence (AI) in the diagnosis of coeliac disease, from the national patient and carer survey (n=231). **(A)** Initial sources of AI awareness; **(B)** general attitude towards AI application in daily life; **(C)** patient acceptance of a clinician’s use of AI-assisted diagnosis; **(D)** the necessity of patient awareness of AI-mediated diagnoses; **(E)** the requirement for explicit patient permission for the use of AI in diagnosis; and **(F)** levels of trust in AI diagnostic systems approved by regulatory bodies.

Exploratory analysis of the Coeliac UK national patient and carer survey identified three demographic associations, which survived Benjamini–Hochberg correction for multiple comparisons, but showed limited effect sizes (ε²=0.05–0.10; Supplementary Table 3). Respondents with higher educational attainment were more likely to have first heard about AI via a scientific publication or forum, rather than the news (18–20% of postgraduate and doctorate holders compared with 0–3% of those with school-level education only, adjusted *p*=0.006), and less likely to consider patient permission necessary before AI use (25.0% of doctorate holders compared with 77.8% of those with no formal education; adjusted *p=*0.022). Respondents with higher levels of education also reported progressively greater trust in AI-assisted diagnosis (e.g. ‘very strong’ trust reported by 11.1% of those with no formal education compared with 37.5% of those with a doctorate; Spearman ρ=0.20, adjusted *p=*0.022). There were no significant associations with gender or age.

### Patient and carer focus groups

Across all four focus groups, participants valued objectivity, consistency and the prospect of faster, more accurate diagnosis. They would accept AI if it had regulatory approval, was more accurate than current practice, and was subject to some form of human-mediated quality control (e.g. batch review, review of uncertain cases). Patients discussed how they already trusted machine-mediated tests in other contexts, with one participant commenting: “We trust lots of health tests which use machines and tests that aren’t 100% accurate, like blood tests even” (Supplementary Table 4). For AI-based diagnosis, they wished that the resulting clinical diagnostic report might be written in plain language they could understand, reflecting a broader frustration that current reports, written for clinicians, are often inaccessible to patients. Some were unaware that other tests were already automated, and were unconcerned to learn that their own past results had been generated without human input. Terminology mattered throughout and was reinforced strongly in our Cambridge session: “artificial intelligence” was often seen as off-putting or associated with less reliable chatbots^57^, whereas “machine learning” was seen as narrower and more clinically trustworthy. Cambridge participants explicitly distinguished this validated, narrow-purpose system from general-purpose large language models (LLMs), and felt the distinction should be made explicit in patient materials (Supplementary Table 9).

Patients’ reservations included loss of human oversight, lack of explainability of “black-box” decisions, cybersecurity, false negatives, and the risk that narrowly trained AI could miss other conditions. Patients wanted clear protocols for human review and for validating software updates. Many emphasised that delays in the diagnosis of CD (which does not necessarily apply to any other duodenal biopsy diagnoses) largely arise pre-biopsy. These include primary care misdiagnosis of CD as anaemia or irritable bowel syndrome and struggles to access endoscopy, with one participant describing a 20-year gap between symptom onset and diagnosis. Participants felt that AI’s promise should be communicated around accuracy and consistency rather than implied improvements in biopsy reporting time, since, for those ultimately diagnosed with CD, the major bottleneck often lies elsewhere. Some had strong concerns about demographic inclusivity, reflecting awareness that training-data quality and biopsy variation across age, ethnicity and diet could affect representativeness. Others raised AI’s environmental cost, seeking reassurance that energy use would be measured and weighed against efficiency gains. Several also highlighted difficulty sharing data between NHS trusts, leading to duplications in testing, with one participant describing an unnecessary repeat duodenal biopsy because a prior diagnosis in another trust was not visible outside that Trust (Table 2). They questioned whether similar problems might limit the training and evaluation of AI at scale.

**Table 2.** Qualitative themes emerging from patient focus groups. Themes and subthemes were identified by inductive thematic analysis of free-text responses and discussion notes from the four focus-group workshops (events 2–5, Table 1). The right-hand column gives illustrative quotations (in quotation marks) or paraphrased participant views. Abbreviations: AI, artificial intelligence; CD, coeliac disease; NHS, National Health Service.

| **Theme** | ***Subtheme*** | **Evidence (quote or paraphrase)** |
|---|---|---|
| **Acceptance contingent on performance & oversight** | *Accuracy over “who”* | “Why choose a less accurate diagnosis (human) if more accurate options (AI) are available?” |
|  | *Formal assurance* | Would accept AI if approved, with quality assurance/controls and active review of parameters. |
|  | *Human back-stop* | Preference for periodic human checks (e.g., every 30 slides) to reassure users. |
| **Transparency & consent** | *Informed* | Majority would want to know if AI was used in their diagnosis. |
|  | *Necessity of understanding* | Some want simple information about how the system works; others deem such information unnecessary if quality/ regulatory standards are met. |
| **Perceived benefits** | *Objectivity & consistency* | “Machines are objective, humans are subjective”; reduces variation between pathologists. |
|  | *Potential time saving* | Could free pathologist time and triage backlogs, might improve outcomes for other patients with high mortality conditions like cancers. Recognition that clinical pathway bottlenecks in CD diagnosis often occur at an earlier stage. |
| **Reservations & risks** | *Loss of human oversight* | “I would want a human to check my sample.” |
|  | *Updates and longevity* | Concern about update control across centres; desire to know how software updates are issued/validated. |
|  | *Missing other conditions* | Want reassurance other conditions won’t be missed if pathway is focused too narrowly on CD. |
|  | *Data governance* | Concern about non-UK ownership of AI systems and third-party access to data (e.g. citing large international technology providers by way of contrast) |
|  | *Terminology* | “Artificial intelligence” perceived as more off-putting and less understandable than “machine learning”, and generalist chatbot tools felt qualitatively different from a narrow, validated medical model. |
|  | *Sustainability* | A minority raised the environmental/carbon cost of AI more broadly and asked whether energy use would be reported and offset by efficiency gains elsewhere in the pathway. |
| **Contextual limitations** | *Delays are often pre-biopsy* | Many delays stem from waiting times for endoscopy/biopsy procedure, not biopsy report speed. |
|  | *System fragmentation* | Frustration that data cannot easily be shared between NHS trusts, complicating both individual care coordination and future training/evaluation of AI tools. |

During the session in Bury St Edmunds, in response to the open questions “Who needs convincing that the method works?”, “How might we convince them?” and “Perceived risks” (Supplementary Table 5), participants identified a wide range of audiences needing persuasion to permit adoption of the technology (patients and families, pathologists and other healthcare professionals, NHS commissioners and regulators, funders, the public). They favoured clear and available evidence of accuracy, transparent validation reporting, and engagement with media, charities and policymakers. They also raised early concerns about training-data diversity, regulatory approval, cybersecurity, and the risk that cost saving, rather than patient benefit, could be perceived as the primary motive for adoption. Similar themes were raised by patient and carer participants in Cambridge, and by healthcare professionals (below).

Patient and carer discussion in Edinburgh (Supplementary Table 6) raised equity/ inclusivity considerations at two levels: firstly, in diagnostic and research practice using AI (e.g. gender bias, with anaemia in women being attributed to periods and not investigated further; “too much research based on men”). There was a concern that a system could be “trained on limited populations” and risk “missing older women / minority ethnic groups”, with the need to “include different races in training data”. Observations about fragmented data-sharing between primary and secondary care were also raised as a barrier to equity/ inclusivity. Secondly, concerns relating more specifically to the acceptance of AI specifically by particular groups were raised. One participant, despite two prior GP misdiagnoses, said they would “give AI a go”. Two participants offered differing generational views on “machine learning” versus “AI” terminology, both preferring the former terminology, with the younger generation being more comfortable with machine learning than the older generation.

At the patients’ and carers’ focus group sessions in Cambridge, where educational attainment was notably high (79.4% educated to degree level or above; Supplementary Table 8), we examined views before and after an educational session focusing on the proposed AI-based technology for duodenal biopsy diagnosis and its safeguards. To assess changes in patients’ views, we asked all participants the same set of questions at the start and end of the session (Supplementary Figures 5-6; results in Table 3). Trust in AI in healthcare (*p=*0.003), belief that AI could improve duodenal biopsy diagnosis (*p<*0.001) and acceptance of AI-assisted diagnosis (*p=*0.001) were significantly higher after the session. Participants’ general opinions of AI became more positive and the perceived importance of explainability increased, neither reaching statistical significance (*p=*0.122 and *p=*0.059, respectively). No views relating to AI generally or in diagnosis became more negative after the educational session. Participants’ views of AI in general and in healthcare had changed also differed significantly (*p*=0.006 and *p*<0.001 respectively; Supplementary Table 8). Pre-session, these questions referred to change over the preceding two years, whereas the post-session questions referred to change by the end of the workshop, so they are not directly comparable.

**Table 3.** Attitudes to AI before and after the Cambridge patient and carers’ workshop explaining the technology and its safeguards. The first five statements were rated for agreement on a five-point scale (1, strongly disagree; 2, disagree; 3, neither agree nor disagree; 4, agree; 5, strongly agree); n=34 patients and carers, with matched pairs of “before” and “after” responses. For the last two statements, participants rated how their view had changed (1, much more negative; 2, slightly more negative; 3, stayed the same; 4, slightly more positive; 5, much more positive): over the previous two years in the “before” ratings, and by the end of the workshop in the “after” ratings. Medians and means are of these 1–5 scores. The percentage columns show the proportion scoring 4 or 5. “Mean increase” is the mean within-participant change in score, tested with the Wilcoxon signed-rank test. See also Supplementary Figures 5–7 and Supplementary Tables 8–9. P-values are unadjusted; significant results remained significant after Bonferroni correction (α≈0.0071). Abbreviations: AI, artificial intelligence; ns, not significant.

| **Attitude statement** | **Median (mean) Likert-scale score before workshop (n=34)** | **Before workshop: Percentage who agree/ strongly agree (rows 1-5), or whose attitude has become slightly/ much more positive (rows 6-7)** | **Median (mean) Likert-scale score after workshop (n=34)** | **After workshop: Percentage who agree/ strongly agree (rows 1-5), or whose attitude has become slightly/ much more positive (rows 6-7)** | **Mean increase (Wilcoxon signed-rank test; ns = not significant)** |
|---|---|---|---|---|---|
| **I feel positively about AI in general** | 3.0 (3.12/5) | 41.2% | 3.5 (3.35/5) | 50.0% | 0.24, *p*=0.122 (ns) |
| **I trust the use of AI in healthcare** | 3.0 (3.32/5) | 38.2% | 4.0 (3.76/5) | 67.6% | 0.44, *p*=0.003 |
| **AI could improve the diagnosis of coeliac disease** | 4.0 (3.62/5) | 52.9% | 5.0 (4.56/5) | 97.1% | 0.94, *p*<0.001 |
| **I would feel comfortable if AI assisted doctors in diagnosing coeliac disease** | 4.0 (4.03/5) | 85.3% | 4.5 (4.47/5) | 97.1% | 0.44, *p*=0.001 |
| **It is important that medical AI systems can explain how they make decisions** | 5.0 (4.50/5) | 94.1% | 5.0 (4.74/5) | 100% | 0.24, *p*=0.059 (ns) |
| **Over the past 2 years, my view of artificial intelligence has become…** | 3.5 (3.21/5) | 50% | 4.0 (3.85/5) | 70.6% | 0.65, *p*=0.006 |
| **Over the past 2 years, my view of artificial intelligence in the delivery of healthcare has become…** | 4 (3.71/5) | 64.7% | 4.0 (4.38/5) | 94.1% | 0.68, *p*<0.001 |

During breakout discussions (Supplementary Tables 7 and 9), participants identified three factors shaping their views: clarity about how the AI was trained and validated, understanding of the safeguards and human checks that would remain in use, and details of its performance relative to pathologists. They contrasted this narrow, regulated application of AI favourably with more sceptical views of generative AI or LLMs. Several participants commented that the session’s use of the terminology “machine learning”, rather than “artificial intelligence” or “AI” was what had changed their view. Recurring concerns included cybersecurity, data integrity, demographic bias in training data, deskilling, and continued human sign-out and post-deployment checks. Several participants requested further public engagement and clearer communication distinguishing this very specific application from generic AI tools.

### Healthcare professionals’ online workshop

The healthcare professional workshop was held online on 7 June 2024, following a brief project overview from Lyzeum Ltd, and was facilitated by “IfM Engage”, a knowledge transfer company within the Institute for Manufacturing (IfM), which is part of the Department of Engineering at the University of Cambridge. Healthcare professionals in attendance represented gastroenterology, histopathology and NHS management. Participants (n=7) were divided into two breakout groups, each first ranking a list of candidate barriers to adoption by vote, then undertaking a structured evaluation of their two highest-ranked barriers (Supplementary Table 12).

The workshop structured barriers to adoption of AI in histopathological diagnosis into four domains:

1. trust/explainability (target populations, sign-out accountability, validation/robustness reporting);
2. privacy/security and hosting (cloud vs. on-premises trade-offs, need for de-identification and data governance);
3. integration and digital infrastructure (heterogeneous NHS laboratory information management systems (LIMS), need for seamless report transfer); and
4. usability/deskilling (concern that AI-assumed cases could de-skill trainees, desire for national training guidance and staged AI exposure).

Proposed implementation strategies included multi-centre testing, departmental validation processes aligned to standardised laboratory accreditation systems such as the UK Accreditation Service (UKAS)^58^, and communication campaigns for clinicians, information technology (IT) departments and patients, the latter being particularly important given the views expressed by patients about wishing to be informed about the use of AI for diagnosis.

### Healthcare professionals’ online questionnaire

The online questionnaire, produced and administered by Lyzeum Ltd, was completed by 37 healthcare professionals between May and November 2025, spanning histopathology, dietetics, gastroenterology and general practice (Supplementary Table 10). The modal healthcare professional preference was for an AI “pathologist’s assistant” over full automation (81.3% vs. 18.8%, n=16), citing the need for assurance, gradual validation, and the clinical value of assisted quantification (Supplementary Tables 10-11). Healthcare professionals indicated that if AI is assisting, it should deliver meaningful insights that support decisions, i.e., it should be explainable. If fully automating, similarly transparent, auditable outputs remain important. Healthcare professionals reported a typical pathologist time of 2 to 8 minutes per duodenal biopsy, wide variation in scanning practice, and uncertainty about the proportion of cases suitable for digital workflows, suggesting that AI might save time chiefly in triage and signing out normal biopsies, with human reporting remaining essential for most other biopsy diagnoses.

### Comparison between healthcare professionals’ and patients and carers’ perspectives

To consider where patients and carers’ and healthcare professionals’ views aligned or diverged, we synthesised recurring themes across both evidence streams, comparing the prevailing views between them across five themes (Table 4): role for AI, transparency and consent, perceived risks, where AI adds most value, and the effect of explanation/education. Both groups agreed AI could save time and improve accuracy. Their views converged on the need for transparency about what AI was being used for and the evidence of its appropriate performance for that task. Healthcare professionals specifically articulated a desire for an AI-assistant as a first approach, whereas patients were less specific, but wanted retention of some form of human oversight. Differing, but complementary sets of risks were identified by the two groups, both requiring effective governance.

**Table 4.** Comparing perspectives in overlapping themes. Themes were synthesised by triangulating the national survey, focus groups, healthcare professional questionnaire and workshop (Table 1). The final column summarises where views converged or differed. Abbreviations: AI, artificial intelligence; CD, coeliac disease.

| Theme | Patients and carers | Healthcare professionals | Net view |
|---|---|---|---|
| **Role for AI** | Accept<br>AI if validated; want human oversight/ review where needed. | Prefer AI assistant initially; automation only after robust evidence. | Similar views. Healthcare professionals specifically articulated a desire for an AI assistant as a first approach, whereas patients were less specific, but wanted retention of some form of human oversight. |
| **Transparency** | Prefer to be told AI was used; mixed<br>on permission and consent. | Support transparency; consent varies between clinical and research use. | Converge on openness; concerns may reduce once AI-mediated diagnostic practice becomes routine. |
| **Risks** | Mechanistic understanding, false negatives, security, data governance and terminology | Deskilling of consultant staff, difficulties training trainee staff, integration burden, variable compliance, hosting risks. | Differing, but complementary sets of risks identified by the two groups; both require effective governance. |
| **Where AI can help** | Objectivity, consistency; triage backlogs (but delays, at least in CD diagnosis, often pre-biopsy). | Triage, quantification of particular parameters, backlog relief if integrated with relevant systems. | Agreement on potential time-saving and improvements in accuracy. |
| **Effect of explanation/education** | Acceptance of, and trust in, AI were higher after a single structured explanation of methodology and safeguards (Cambridge) | Emphasise that explainability of diagnostic decisions, ongoing validation, audit and clear communication are central to earning and sustaining trust. | Converge on transparent, evidence-led communication as a precondition for acceptance by all stakeholders. |

## Discussion

To our knowledge, this is the first study to combine patient, carer and healthcare professional perspectives on AI-assisted duodenal biopsy diagnosis. Its strengths include the triangulation of a national-level patient survey data with focus-group narratives and healthcare professionals’ inputs from both a dedicated survey and a structured workshop, giving a multi-perspective view of acceptability, governance expectations and implementation realities.

### Common themes across stakeholder groups

Our synthesis suggests overall cautious support for the use of AI in duodenal biopsy diagnosis, contingent upon evidence of accuracy and time-saving, transparency about the purposes for which the AI is being used and robust governance, in terms of human oversight of the diagnostic process and broad risk mitigation (Table 4). Most patients wanted to be informed when AI was used, but only about half felt permission necessary, often likening validated AI to other laboratory technologies where a clinician remains accountable. Healthcare professionals preferred an “AI-assistant” model over full automation, while patients wanted some human involvement to be retained, perhaps in the form of spot-checks/ audit, and human intervention in all cases of uncertainty. Both groups prioritised intelligibility and accountability over technological novelty. Triangulating patient and carer and healthcare professional data points suggests that an acceptable common starting model for both groups might be either AI quantifying parameters and highlighting features (e.g., with heat-maps) and providing a suggested diagnosis with confidence scores, alongside human sign-out, although some patients and carers would also accept full automation with spot-check quality control.

The term “machine learning” understood as a description of a narrow tool for a specific clinical task, was consistently distinguished from “artificial intelligence” in its broader, generalist sense across all four patient focus groups, the latter attracting more scepticism. Participants felt this distinction is rarely made in public discourse, and that making it explicit in patient-facing materials could reduce unwarranted anxiety without overstating the technology’s capabilities. This finding arose from a discussion setting rather than a tested communication intervention and should be interpreted with caution, but it does echo previous studies indicating that terminology shapes trust in algorithmic systems^59 60^.

Patients and carers’ and healthcare professionals’ views converged on where AI’s practical value lay, and the infrastructure required. Both groups identified backlog reduction via AI-assisted triage as a potential benefit, though CD patients cautioned that delays in CD diagnosis (but not necessarily in other conditions diagnosed on the basis of a duodenal biopsy) often arise pre-biopsy, in primary care or in difficulties accessing an endoscopy, rather than due to slow diagnostic biopsy reporting. Healthcare professionals were very clear that successful implementation was contingent upon practical infrastructure (LIMS integration, cross-institutional interoperability, and hosting decisions). A minority of participants raised ethical considerations about equity and inclusivity, as well as some being concerned about AI’s environmental footprint and fragmented NHS data-sharing.

### Differences between views of stakeholder groups

Divergence between the stakeholder groups most likely represented differing expertise and experience. Healthcare professionals’ caution about full automation centred on specific, technical prerequisites, including the need for detailed validation datasets, audit trails and regulatory sign-off. Only a minority (18.8%, 3/16) preferred full automation, citing reduced review burden and clearer accountability, while patients’ appetite for full automation was less clear-cut and not consistently measured across our sessions. As one might expect, patients’ key concern was retaining a human point of contact with whom to discuss their diagnosis, rather than having particularly specific views on how the diagnosis should be made. Healthcare professionals raised an important concern that AI-mediated diagnosis of routine cases could reduce trainees’ and consultants’ exposure to straightforward biopsies, making both training, and retention of a view of the normal range of histological appearances, difficult. Using audit/ oversight processes of the AI method as an opportunity to look at small numbers of normal biopsies was briefly discussed. Inevitably, the views of the patient population were not homogeneous. Respondents with higher educational attainment were less likely to deem explicit permission to use AI necessary (Supplementary Table 3), suggesting that communication strategies may need tailoring to educational level, rather than a one-size-fits-all approach.

### Attitudes before and after an educational session

Acceptance of, and trust in, AI increased after an educational session about the AI-based diagnostic approach and its safeguards (Table 3). This supports the value of education and information strategies for patients and the public if AI-based biopsy reporting is introduced. Separately from the educational session, we were unable to reliably distinguish change in public attitudes over the roughly two-year study period from demographic differences (in age, gender and educational level) between the cohorts recruited at each site, and the exact approach to facilitation, as the group of facilitators varied between sessions. However, this would be a fascinating and important topic for future study.

### Limitations

Although our study samples the views of almost 400 duodenal biopsy diagnosis stakeholders, this remains a small sample in comparison with the approximately 400,000 duodenal biopsies performed annually in the UK^61^, risking bias. Participants in our focus groups, workshop and questionnaires all volunteered to take part, so our samples may be prone to selection bias, over-representing those with stronger views, greater research interest, or higher digital literacy or health awareness. If those most averse to AI were the least likely to take part, this would bias our results towards a more favourable stakeholder view of AI. Our focus-group design was geographically biased in that it focused on urban centres in the UK, meaning that its exact relevance to rural UK populations and non-UK populations remains unknown. Within the Coeliac UK national survey, educational level showed some association with respondents’ views (Supplementary Table 3). Our sample is skewed towards more educated and female respondents relative to the general population, meaning that attendees’ views should be interpreted as indicative rather than population-representative. Age, gender and educational level were not consistently recorded across all focus groups, making it harder to determine the likelihood of skewing of participants’ views by these factors.

A more fundamental limitation is that both the national survey and the focus groups were promoted to, and therefore predominantly completed or attended by, Coeliac UK members. While Coeliac UK membership is broadly representative of the diagnosed CD population, it is unlikely to represent the undiagnosed CD population, who are disproportionately more likely to be from socioeconomically deprived backgrounds^62 63^ than Coeliac UK’s membership, which is skewed towards less deprived individuals. Secondly, it is not necessarily representative of the >90% of people undergoing duodenal biopsy who receive a diagnosis other than CD^28^. A related consequence is that most respondents and attendees had already completed their diagnostic journey, meaning that this study does not capture the views of people who are currently undiagnosed or currently undergoing diagnosis, who, arguably, represent the population with the most at stake in how any future AI-assisted pathway is designed.

Our study captures extensive qualitative data, but we recognise that it reflects discussions at a particular point in time, at which particular AI capabilities are understood by the general public and for which regulatory guidance continues to evolve. A similar study undertaken years or even months later is likely to elicit substantially different views in this fast-moving field. Finally, patient and carers’ and healthcare professionals’ data were gathered using different methodologies, administered by different facilitators, potentially causing variation in the views recorded between focus groups or breakout discussion groups within them.

## Conclusions

Patients and healthcare professionals are cautiously supportive of AI-assisted diagnosis of duodenal biopsies, conditional upon demonstration of accuracy, appropriate human oversight and transparent communication. Structured public explanation measurably improves attitudes towards use of AI in diagnosis.

## Data availability

The original contributions presented in this study are included in the article/ supplementary materials. Further inquiries can be directed to the corresponding author.

## Authorship

Conceptualisation and design: All authors

Work with stakeholders: All authors

Data extraction: KS, JB, JW

Data interpretation: All authors

Manuscript drafting: JB, FJ, KS, EJS

Manuscript review: All authors

Supervision: EJS

## Ethical approval

This study is a synthesis of anonymized views expressed at various engagement events. It contains no identifiable patient information, and therefore ethical approval is not required.

## Patient consent

No identifiable patient information is included. Consent to participation in the events and questionnaires was implied by voluntary participation.

