## Supplementary for "Patient, carer and clinical stakeholder views on using artificial intelligence and machine learning for the histopathological diagnosis of duodenal biopsies"

**Supplementary Materials**

Supplementary document assembled from source questionnaires, submitted responses and further analysis.

**Glossary of Terms and Abbreviations**

***Study***

| **Term** | **Definition** |
| --- | --- |
| **AI** | Artificial intelligence |
| **CD** | Coeliac disease |
| **HCP** | Healthcare professional |
| **Lyzeum** | The AI-based diagnostic system evaluated in this study |
| **PPIE** | Patient and Public Involvement and Engagement |

***Clinical***

| **Term** | **Definition** |
| --- | --- |
| **Duodenal biopsy** | Small-intestinal tissue sample taken via endoscopy and examined by a pathologist to diagnose CD |
| **IBS** | Irritable bowel syndrome; a common alternative or delaying diagnosis |
| **Marsh score** | Histological grading scale for the severity of coeliac-related mucosal damage |
| **tTG** | Tissue transglutaminase; a blood antibody test used to screen for CD |
| **Villi / crypts** | Microscopic small-intestinal structures assessed by pathologists; CD causes villous atrophy and crypt hyperplasia |

***Statistics (Supplementary Table 3 and elsewhere)***

| **Term** | **Definition** |
| --- | --- |
| **BH correction** | Benjamini-Hochberg procedure controlling the false discovery rate across multiple comparisons |
| **Effect size (ε², ρ)** | Standardised strength of an association, independent of sample size |
| **Kruskal-Wallis** | Non-parametric test comparing a variable across three or more groups |
| **Spearman’s ρ** | Non-parametric, rank-based correlation coefficient |
| **Wilcoxon signed-rank** | Non-parametric test comparing paired (matched pre/post) measurements |

***Education levels***

| ***Term*** | ***Definition*** |
| --- | --- |
| ***GCSE / BTEC / PGCE*** | *UK qualifications: secondary school; vocational (Business and Technology Education Council); teaching (Postgraduate Certificate in Education)* |

### Supplementary Methods: Data Collection Instruments

The following Supplementary Figures reproduce the questionnaires, discussion guides and feedback forms used across the seven data-collection activities described in Methods (Table 1).

#### Supplementary Figure 1

**National online patient and carer questionnaire (Coeliac UK, February 2024 – December 2025)**

**Intro. “Use of AI to diagnose coeliac disease.” A group of researchers in the UK are developing an artificial intelligence system to help improve the speed and accuracy of diagnosing coeliac disease. Artificial intelligence can be described as a computer system that uses information to learn and to recognise patterns independently, without human interference. This questionnaire is intended for people aged 16 years and over, with or without a current diagnosis of coeliac disease. All results are collated and reported anonymously.**

**Q1. Before today had you ever heard of artificial intelligence and if so how did you FIRST hear about it?**

☐ Never heard of it before today

☐ Friends or relatives

☐ News

☐ A scientific publication or forum

☐ A science fiction book/movie/series

☐ Other (please specify)

**Q2. How do you feel about the application of artificial intelligence in day-to-day life? [Any comments free text]**

☐ Very positive

☐ Positive

☐ Neutral

☐ Negative

☐ Very negative

**Q3. Would you accept a healthcare professional relying on the use of artificial intelligence to diagnose coeliac disease? [Any comments free text]**

☐ Yes

☐ No

☐ Unsure

**Q4. Do you feel it's necessary for a patient to be AWARE that their diagnosis of coeliac disease was made using an artificial intelligence system? [Any comments free text]**

☐ Yes

☐ No

☐ Unsure

**Q5. Do you feel it's necessary for a patient to be asked for their PERMISSION to use an artificial intelligence system in the diagnosis of their coeliac disease? [Any comments free text]**

☐ Yes

☐ No

☐ Unsure

**Q6. What level of trust would you have in a diagnosis of coeliac disease that had been made using an artificial intelligence system that had been approved by all the necessary regulatory bodies? [Any comments free text]**

☐ Very strong

☐ Strong

☐ Moderate

☐ Weak

☐ Very weak

☐ None at all

☐ Unsure

**Q7. Approximately how long have you been diagnosed with coeliac disease?**

☐ Not diagnosed – Less than 1 year – 1 to 10+ years (annual bands) – Not sure

**Q8. Do you have any immediate family members with a diagnosis of coeliac disease? (tick all that apply)**

☐ Parent

☐ Sibling

☐ Child

☐ None of the above

**Q9. What is your age? (years, free entry)**

**Q10. What is your gender?**

☐ Male

☐ Female

☐ Prefer not to say

☐ Other (please specify)

**Q11. What is your highest level of education?**

☐ No formal education

☐ GCSE / O level

☐ A level / BTEC

☐ Undergraduate degree

☐ Postgraduate degree

☐ Doctorate

**Q12. [Registration of interest in the Stevenage focus group – name / email / location. Omitted here in full; see Methods for aggregate participation figures (69/231 registered interest).]**

#### Supplementary Figure 2

**Focus-group discussion guide - Stevenage (March 2024)**

**Session. Coeliac UK Stevenage Group, March 2024**

**BREAKOUT 1**

***Points to consider***

**– For your diagnosis of coeliac disease (or any other diagnosis) did you ever question or doubt the results?**

**– Did you ever question or ask about the accuracy of the results?**

**– Can you remember how long you had to wait for the results, and did you think this reasonable?**

**BREAKOUT 2**

***Points to consider***

**– What, if anything, would you like the researchers to consider while carrying out this research project?**

**– What human input would you expect in the process?**

**– Under what conditions would you be happy for AI to be included within the diagnosis process?**

**– Are you aware of any other health conditions currently using AI technology, if so which ones?**

#### Supplementary Figure 3

**Focus-group discussion guide - Bury St Edmunds (June 2025)**

**Session. Coeliac UK Bury St Edmunds Group, 14th June 2025**

**1. In your view(s), why does it take so long to diagnose coeliac disease?**

**2. [“Oak tree” sorting exercise] Oak tree numbers in group 1 / group 2; what features did you use to divide them up?**

**3. Who do we need to convince that our method works?**

**4. How might we do that?**

**5. Where are the potholes? Your thoughts on risks to our strategy and any suggested improvements.**

#### Supplementary Figure 4

**Focus-group discussion guide - Edinburgh (September 2025)**

**Session. Coeliac UK Edinburgh Group, 26th September 2025**

**1. In your view(s), why does it take so long to diagnose coeliac disease?**

**2. [“Oak tree” sorting exercise] Oak tree numbers in group 1 / group 2; what features did you use to divide them up?**

**3. Who do we need to convince that our method works?**

**4. How might we do that?**

**5. Where are the potholes? Your thoughts on risks to our strategy and any suggested improvements.**

#### Supplementary Figure 5

**Cambridge PPIE workshop - Questionnaire 1 (pre-session, June 2026)**

**Q1. What is your age (years)?**

**Q2. What is your gender?**

☐ Male

☐ Female

☐ Prefer not to say

☐ Other (please specify)

**Q3. Education**

☐ Left school at 16

☐ Left school at 18

☐ Undergraduate degree

☐ Postgraduate degree (please specify)

**Q4. Yes/No: (a) Have you/a family member been diagnosed with coeliac disease? (b) Have you ever had an upper GI biopsy taken? (c) Have you ever worked in healthcare? (d) Have you ever worked in technology or AI?**

**Q5. Likert agreement (Strongly agree – Strongly disagree) with: ‘I feel positively about AI in general’; ‘I trust the use of AI in healthcare’; ‘AI could improve the diagnosis of coeliac disease’; ‘I would feel comfortable if AI assisted doctors in diagnosing coeliac disease’; ‘It is important that medical AI systems can explain how they make decisions’**

**Q6. Over the past 2 years, my view of AI in general / in the delivery of healthcare has become… (Much more positive – Much more negative)**

#### Supplementary Figure 6

**Cambridge PPIE workshop - Questionnaire 2 (post-session, June 2026)**

**Q1. Same five Likert statements as Questionnaire 1, Q5, re-administered post-session.**

**Q2. At the end of this workshop, my view of AI in general / in the delivery of healthcare has become… (Much more positive – Much more negative)**

**Q3–5. Free text: what (if anything) changed your view; any remaining questions or concerns.**

**Q6. Workshop evaluation (Strongly agree – Strongly disagree): ‘The workshop was well organised’; ‘I enjoyed attending the workshop’; ‘I felt my voice was heard and my contribution was valued’; plus free text comments.**

#### Supplementary Figure 7

**Focus-group discussion guide - Cambridge (June 2026)**

**Session. Cambridge PPIE Workshop, Hughes Hall, June 2026**

**1. How do you use AI in daily life? If you don’t use it, what stops you from using it?**

**2. How do you think AI might be useful in healthcare? Do you have any concerns around the use of AI in healthcare?**

**3. How would you feel about a regulated programme which lets you put your symptoms in and give you a diagnosis?**

**4. How do you think people might react if a computer system gave a different diagnosis to one given by a trained histopathologist in the past? (i.e., retrospective)**

`

#### Supplementary Figure 8

**Healthcare professional online questionnaire (Lyzeum Ltd, May–November 2025)**

**Demographics:**

**1. Current role: gastroenterologist / histopathologist / GP / nurse endoscopist / dietitian / other (free text)**

**About your centre:**

**2. In your centre, are duodenal biopsies placed in the same block and on the same slide as biopsies from other GI tract sites (e.g., gastric biopsies)?**

☐ Yes

☐ No

**3. Roughly what proportion of upper GI endoscopic biopsy cases (1 case = 1 set of patient samples) contain duodenum only?**

☐ 0% of cases

☐ 10% of cases

☐ 20% of cases

☐ 30% of cases

☐ 40% of cases

☐ 50% of cases

☐ 60% of cases

☐ 70% of cases

☐ 80% of cases

☐ 90% of cases

☐ 100% of cases

**4. In your centre, would it be feasible to have a "duodenal endoscopy looks normal" box either in the endoscopy report, or on the pathology request form accompanying the biopsies?**

☐ Yes for both

☐ Yes, only for endoscopy report

☐ Yes, only for pathology report

☐ No

**About biopsy reporting:**

**5. In your view, how important is it that a written pathology report describes the microscopic findings in detail, rather than simply stating active coeliac disease?**

**6. In your opinion, would the following text suffice for a duodenal biopsy report? "AI-mediated examination of a digital scan of this biopsy slide has diagnosed active coeliac disease."**

☐ Yes

☐ No

☐ Don’t know

**About your workflow:**

**7. How long on average do you think a pathologist spends in total on each duodenal biopsy, from looking at the slide to completing and signing out the**

**pathology report?**

☐ <2 min

☐ 2–5 min

☐ 5–8 min

☐ 8–10 min

☐ 10–15 min

☐ >15 min

**10. Does your centre scan all of your duodenal biopsies?**

☐ Yes

☐ No

☐ Don’t know

**Attitudes towards AI Approaches:**

**11. If an AI approach to duodenal biopsy reporting was introduced to assist the pathologist (and not to sign out biopsy reports independently), how much time per duodenal biopsy would it need to save the pathologist to be worth introducing?**

☐ <2 min

☐ 2–5 min

☐ 5–8 min

☐ 8–10 min

☐ 10–15 min

☐ >15 min

**12. In your opinion, which of the following scenarios would be preferable and why?**

☐ A - Using an AI system that fully automates the reporting of most duodenal biopsies (i.e. normal biopsies and potentially coeliac disease), including stating active coeliac disease.

☐ B - Using an AI system which merely acts as a "pathologist's assistant", and suggests possible active coeliac disease.

**13. Explanation of preference for Scenario A or B (free text)**

**Hypothetical Scenarios:**

**14. To what extent would you be content with an AI system that automatically did any of the following with duodenal biopsies?**

**a. Only reported biopsies that showed features of coeliac disease?**

**b. Only reported biopsies that were normal?**

**c. Reported both normal and coeliac biopsies?**

☐ Very content

☐ Content

☐ Neutral

☐ Not content

☐ Definitely not content

☐ Don’t know

**15. If we provided an AI assistant, rather than software for fully automated reporting of duodenal biopsies, how useful would it be if it provided the following?**

**a. Providing an intraepithelial lymphocyte and enterocyte count. Only reported biopsies that were normal?**

**b. Villous height to crypt depth ratio**

☐ Very useful

☐ Useful

☐ Neutral

☐ Not useful

☐ Definitely not useful

☐ Don’t know

**16. Conversely, if we provided fully automated reporting of duodenal biopsies, how useful would it be if it provided the following?**

**a. Providing an intraepithelial lymphocyte and enterocyte count. Only reported biopsies that were normal?**

**b. Villous height to crypt depth ratio**

☐ Very useful

☐ Useful

☐ Neutral

☐ Not useful

☐ Definitely not useful

☐ Don’t know

**Benefits and Obstacles to AI Implementation:**

**17. Benefits of AI Implementation for Normal Biopsies:**

☐ Investing in the future

☐ Saving money

☐ Improving objectivity

☐ Saving staff time

☐ Reducing turnaround time

☐ Other (please specify)

**18. Other Benefits: (free text)**

**19. Obstacles of AI Implementation for Normal Biopsies**

☐ Investing in the future

☐ Saving money

☐ Improving objectivity

☐ Saving staff time

☐ Reducing turnaround time

☐ Other (please specify)

**20. Consent to further contact**

#### Supplementary Figure 9

**Healthcare professional online workshop (Lyzeum Ltd, June 2024)**

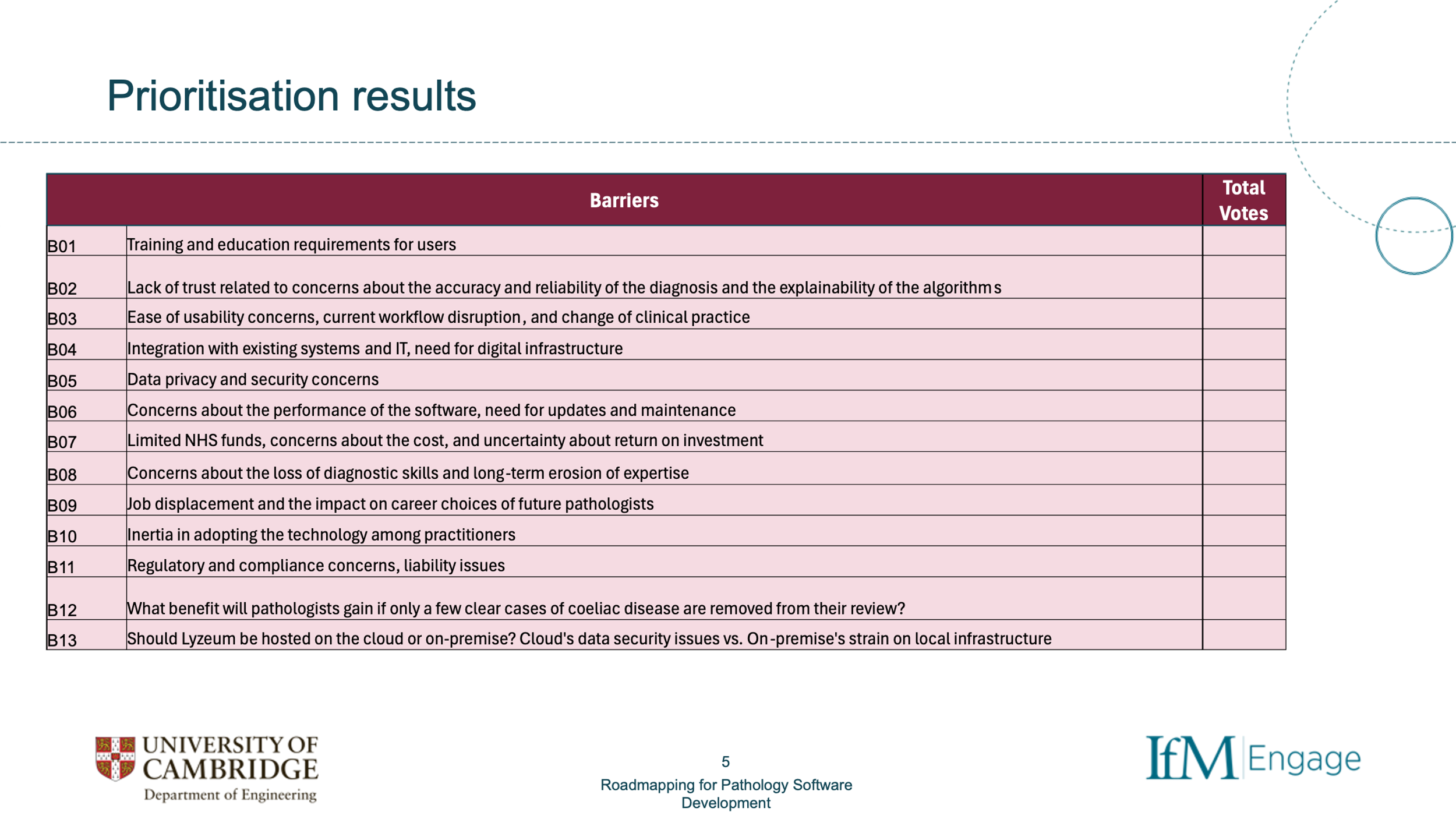

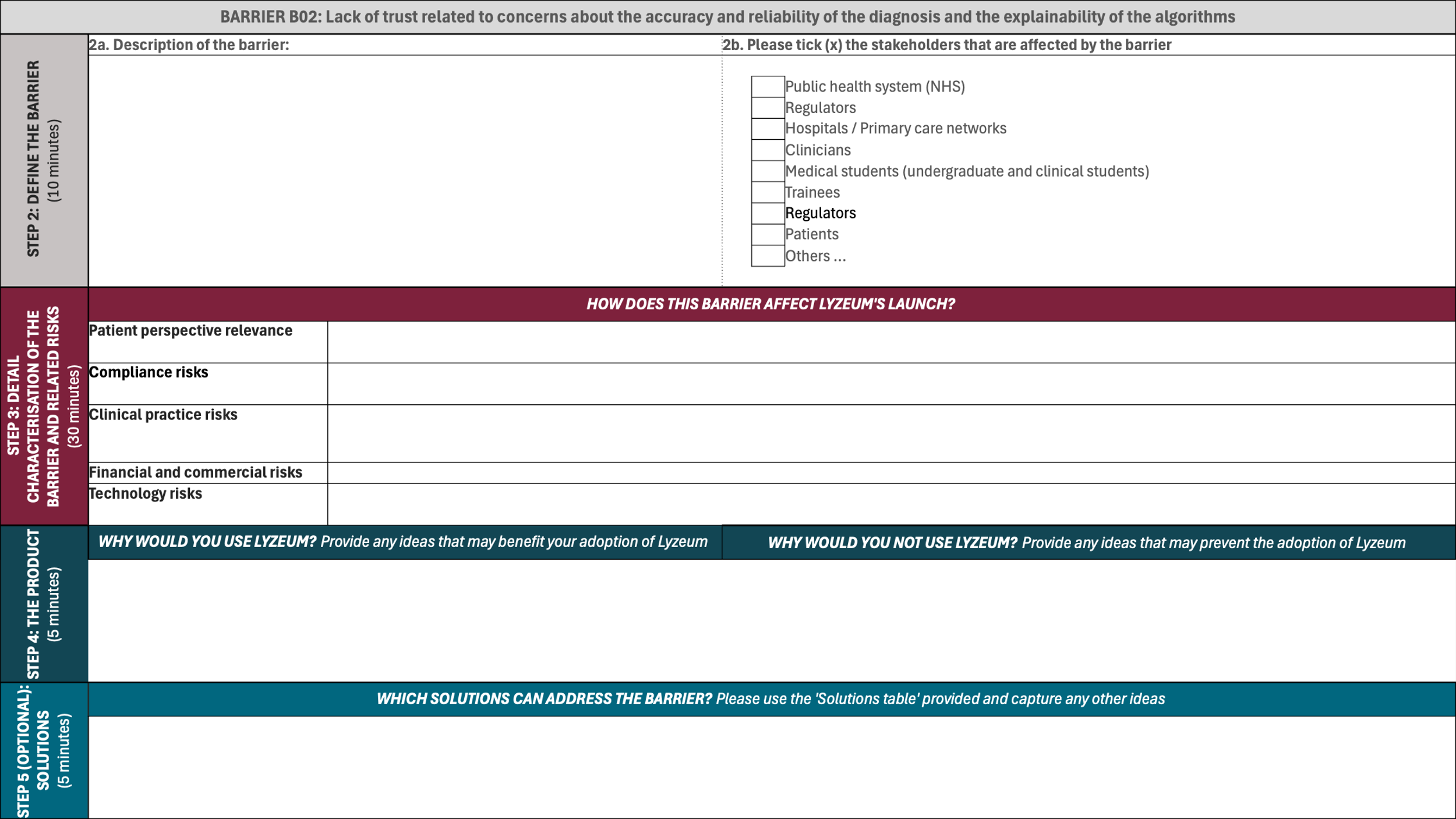

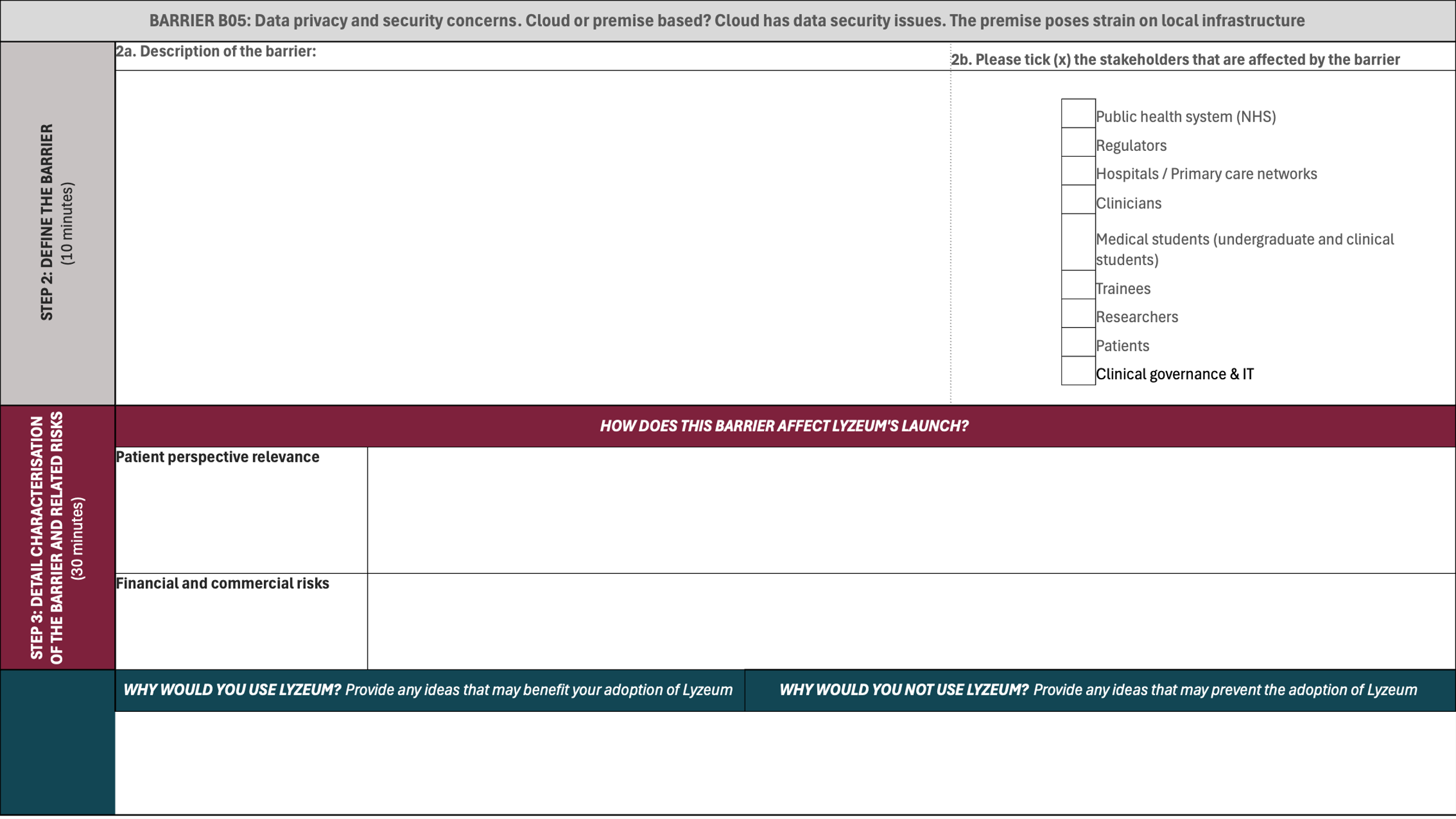

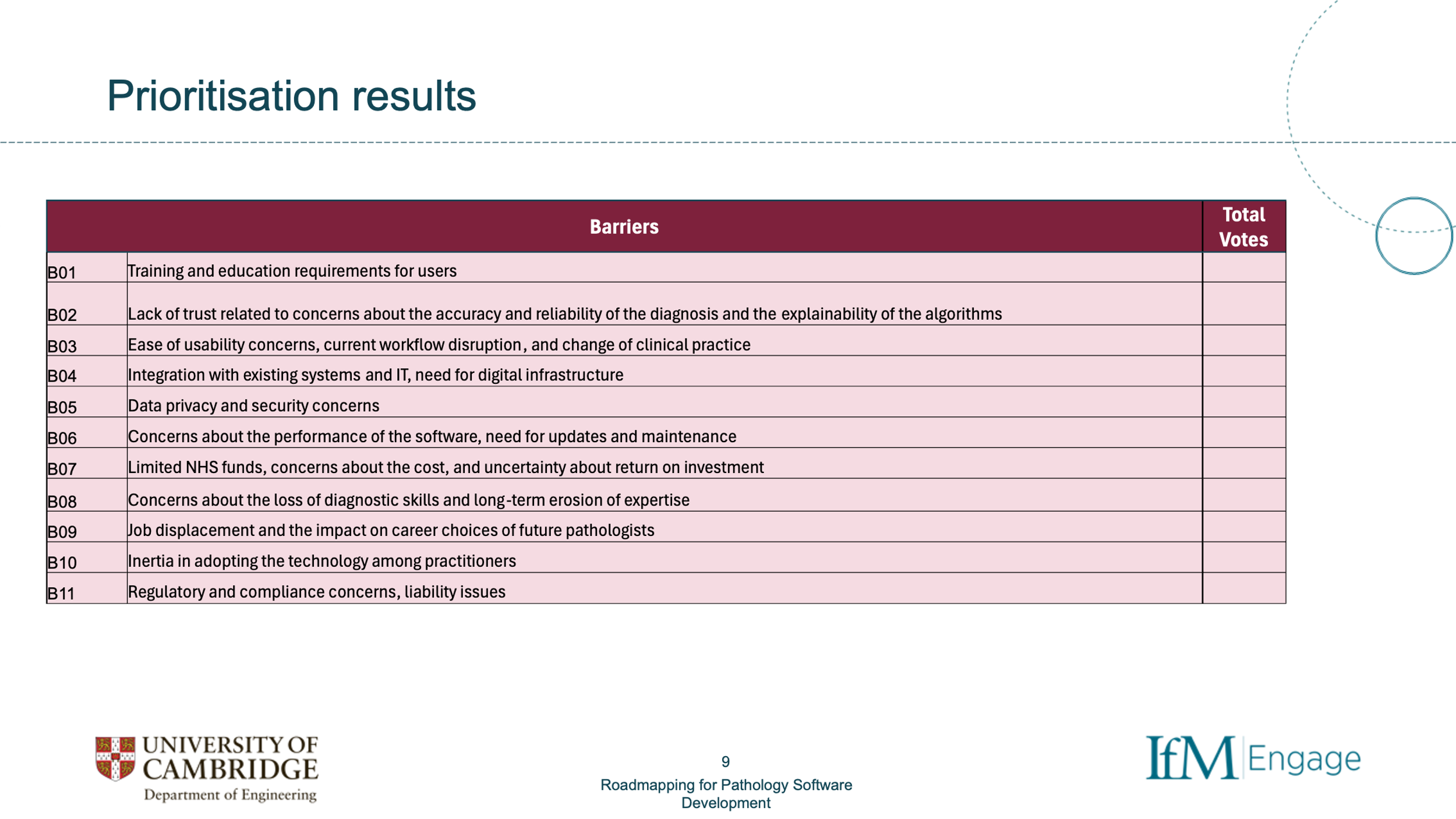

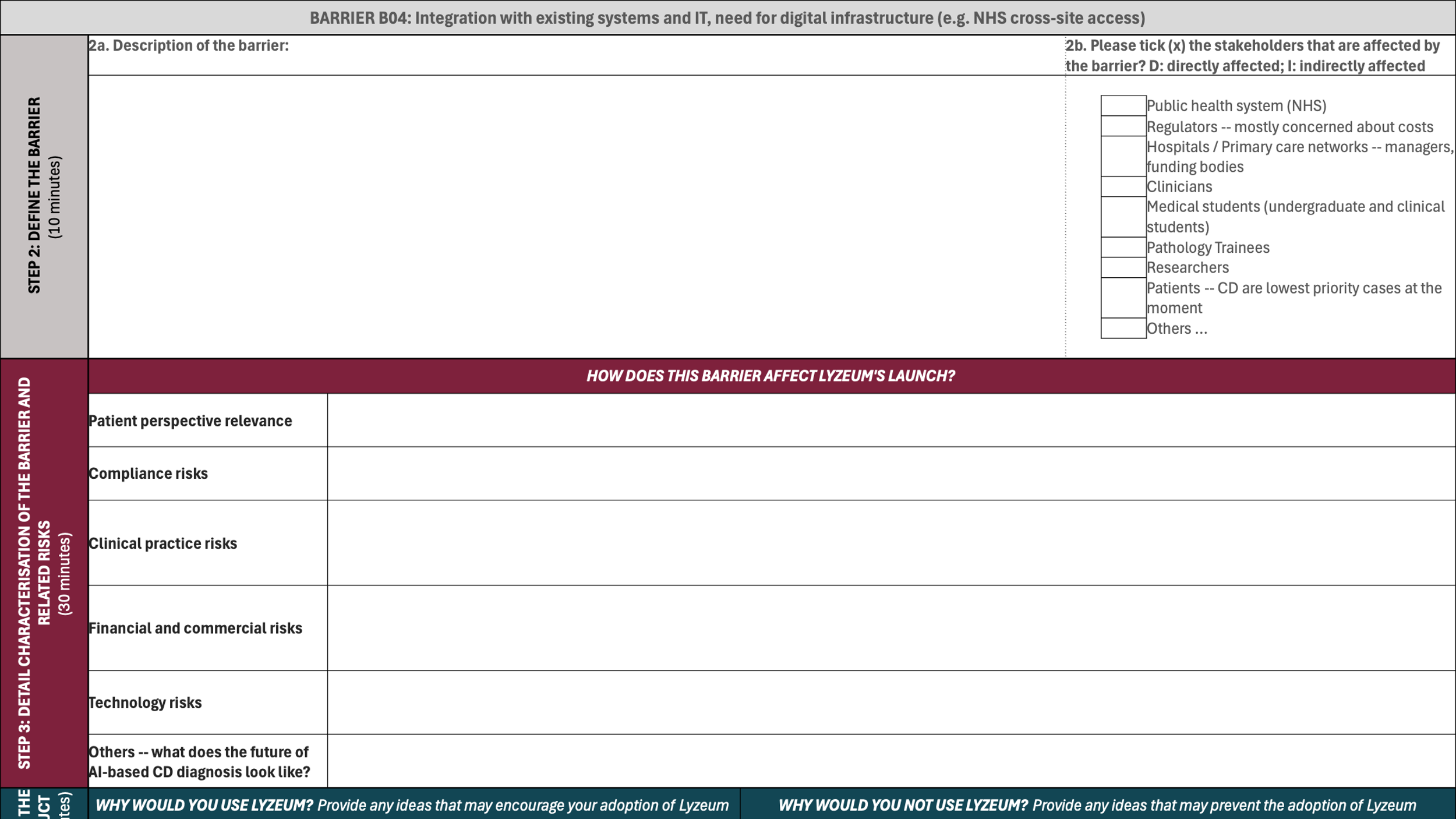

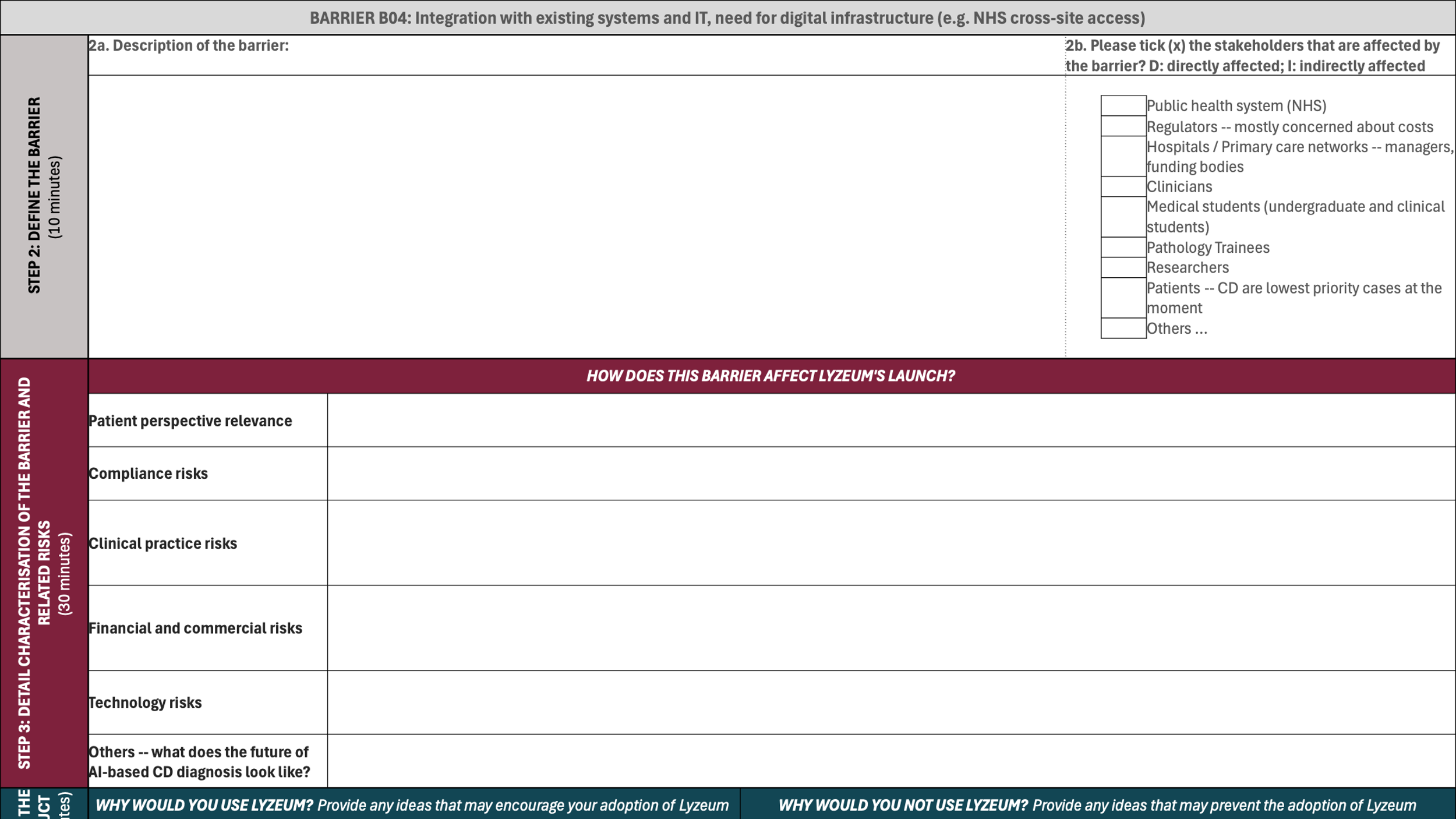

### Supplementary Results: Raw Data

The following Supplementary Tables give the full quantitative and free text breakdown underlying the summary statistics reported in the main Results section.

#### Supplementary Table 1

**National online patient/carer questionnaire - full quantitative results (n=231 unless stated)**

**Q1. Prior awareness of AI**

*Before today had you ever heard of artificial intelligence and if so how did you FIRST hear about it? (please select only ONE option)*

| **Response option** | **n** | **%** |
| --- | --- | --- |
| Never heard of it before today | 12 | 5.2% |
| Friends or relatives | 16 | 6.9% |
| News | 146 | 63.2% |
| A scientific publication or forum | 19 | 8.2% |
| A science fiction book/movie/series | 10 | 4.3% |
| Other (please specify): | 28 | 12.1% |

**Q2. Feelings about AI in day-to-day life**

*How do you feel about the application of artificial intelligence in day-to-day life?*

| **Response option** | **n** | **%** |
| --- | --- | --- |
| Very positive | 36 | 15.6% |
| Positive | 95 | 41.1% |
| Neutral | 72 | 31.2% |
| Negative | 25 | 10.8% |
| Very negative | 3 | 1.3% |

**Q3. Would accept HCP relying on AI to diagnose CD**

*Would you accept a healthcare professional relying on the use of artificial intelligence to diagnose coeliac disease?*

| **Response option** | **n** | **%** |
| --- | --- | --- |
| Yes | 158 | 68.4% |
| No | 13 | 5.6% |
| Unsure | 60 | 26.0% |

**Q4. Necessary for patient to be AWARE AI used**

*Do you feel it's necessary for a patient to be AWARE that their diagnosis of coeliac disease was made using an artificial intelligence system?*

| **Response option** | **n** | **%** |
| --- | --- | --- |
| Yes | 170 | 73.6% |
| No | 37 | 16.0% |
| Unsure | 24 | 10.4% |

**Q5. Necessary for patient PERMISSION for AI use**

*Do you feel it's necessary for a patient to be asked for their PERMISSION to use an artificial intelligence system in the diagnosis of their coeliac disease?*

| **Response option** | **n** | **%** |
| --- | --- | --- |
| Yes | 121 | 52.4% |
| No | 79 | 34.2% |
| Unsure | 31 | 13.4% |

**Q6. Trust in AI diagnosis if approved by regulators**

*What level of trust would you have in a diagnosis of coeliac disease that had been made using an artificial intelligence system that had been approved by all the necessary regulatory bodies?*

| **Response option** | **n** | **%** |
| --- | --- | --- |
| Very strong | 55 | 23.8% |
| Strong | 99 | 42.9% |
| Moderate | 55 | 23.8% |
| Weak | 4 | 1.7% |
| Very weak | 2 | 0.9% |
| None at all | 5 | 2.2% |
| Unsure | 11 | 4.8% |

**Q7. Time since CD diagnosis**

*Approximately how long have you been diagnosed with coeliac disease?*

| **Response option** | **n** | **%** |
| --- | --- | --- |
| I'm not diagnosed with coeliac disease | 20 | 8.7% |
| Less than 1 year | 20 | 8.7% |
| 1 year | 11 | 4.8% |
| 2 years | 14 | 6.1% |
| 3 years | 14 | 6.1% |
| 4 years | 8 | 3.5% |
| 5 years | 6 | 2.6% |
| 6 years | 9 | 3.9% |
| 7 years | 10 | 4.3% |
| 8 years | 3 | 1.3% |
| 9 years | 6 | 2.6% |
| 10 years | 9 | 3.9% |
| More than 10 years | 100 | 43.3% |
| Not sure how long I've been diagnosed | 1 | 0.4% |

**Q8. Immediate family members with CD**

*Do you have any immediate family members with a diagnosis of coeliac disease? (Please tick all that apply)*

| **Response option** | **n** | **%** |
| --- | --- | --- |
| Parent - mother/father | 22 | 9.6% |
| Sibling - sister/brother | 27 | 11.8% |
| Child - son/daughter | 37 | 16.2% |
| None of the above | 152 | 66.7% |

*Note: percentages calculated on n=228 (3 of 231 respondents did not answer this question). This question allowed multiple selections, so response counts sum to more than 228.*

***Q9. Age***

*Free text answers for age: mean 62.9 years, median 65, range 20–91 (n=226 valid numeric responses of 229 answered).*

**Q10. Gender**

*What is your gender?*

| **Response option** | **n** | **%** |
| --- | --- | --- |
| Male | 52 | 22.5% |
| Female | 177 | 76.6% |
| Prefer not to say | 0 | 0.0% |
| Other (please specify): | 2 | 0.9% |

**Q11. Highest level of education**

*What is your highest level of education? Please select the most appropriate or equivalent.*

| **Response option** | **n** | **%** |
| --- | --- | --- |
| No formal education | 9 | 3.9% |
| GCSE / O level | 39 | 17.0% |
| A level / BTEC | 50 | 21.7% |
| Undergraduate degree | 67 | 29.1% |
| Postgraduate degree | 49 | 21.3% |
| Doctorate | 16 | 7.0% |

*Note: percentages calculated on n=230 (1 of 231 respondents did not answer this question).*

#### Supplementary Table 2

**National online patient/carer questionnaire - free text comments by question**

*Comments are reproduced anonymously.*

**Q2. Feelings about AI in day-to-day life (n=64 comments)**

• Depends on what is used for and how secure it is

• Can be used for good or bad

• Where will AI end.

• It is by degree how much it is used. you cannot replace human interaction

• Negative views are in relation to day-today life and the capacity to manipulate people's views

• There are clear benefits and downsides. Evidently more regulation is required

• Just needs to be regulated thoroughly

• To improve efficiency, and effectiveness, improved outcomes then AI is definitely positive, and must be embraced, for instance in medicine.

• Don’t know enough about it to make an informed decision.

• If used by knowledgeable and trained personnel

• Unsure really as don't yet know what the effect will be.

• Only if highly regulated and controlled

• Not sure about that, it should not be taken for granted?

• As long as it is well controlled.

• Negative on private and social life but the exceptions being medical research

• I think there are many good benefits with AI. I do wonder how we can control some aspects of its use.

• As long as care is taken over the diversity of the data that AI learns from I think it can be positive tool

• It has a lot of potential for good, especially in fields such as health and care, but there is a need to address the dangers e.g., data protection, in warfare etc

• I think it has its place, but needs very rigorous safeguards surrounding its use, in particular, in this case, very well trained users

• Some use is positive, some is negative and some is just unreliable

• Particularly in medicine.

• Concerned about scamming

• Could help to sped up a lot of data processing

• I don't mind AI applications. But a human does need to verify results at some point or have regular spot checks to ensure the AI is on track.

• It is a great aid in doing the routine but detailed tasks.

• I think it can be a bit of a fad, especially at the moment, with everyone clambering to make their products "powered by AI" even if it doesn't warrant it.

• So long as it is used in a positive way and is accurate.

• Concerned about the reliability of the technology

• Applied in the right way, AI can be a useful tool across a wide range of applications. Caution needs to be taken to ensure the results are tested and there is a feedback loop to improve accuracy.

• Crucially depends upon context and safeguards

• With controls in some scenarios

• It depends entirely on context. It’s already been shown to be great for certain purposes and rubbish for others.

• Need to be wary, considered

• As long as we don’t become to rely on it

• Positive if not abused and with appropriate regulation

• This could improve diagnosis, and speed up the process to enable lab testing to be carried out quicker.

• Like all tools, it depends on the author/user etc. It needs to be treated with caution.

• Though it is a worry that it might become too clever.

• I find it worrying that it will replace humans in so many jobs. I dislike its use to create algorithms in marketing, internet sites, social media.

• Concern around impact on day to day living. Influence of AI on all areas of life.

• I believe AI has a great potential in healthcare applications

• But with reservations about other applications e.g., deep fake

• If used correctly and safely it can have some fantastic uses.

• If it works for a correct diagnosis, this is a good thing.

• Like all new developments, there are always pluses and minuses

• I would be positive about it in some circumstances but also have reservations about its use

• As always, it all depends on aim, quality, cost, type of application context,.........

• Somewhat concerned but realise there may be benefits

• Some concern it may be used for negative purpose but excited it could be positive in field like productivity and health

• Hopefully used with common sense and as it's investigating a serious condition I would hope an experienced professional would follow up results.

• Hopefully it will be strictly monitored

• My feeling is the value is in how the AI has been trained. Such as the datasets used for training and validation.

• I have a basic mistrust of computers and so would be worried about things like self driving vehicles that could kill someone.

• Positive, with the caveat that AI should avoid biases in design e.g., the "default male" as the model.

• It depends on the particular application

• It’s as good/bad as the initial programmers. Negative because human oversight is so poor. Think brilliant and overdue for medicine.

• I believe it could have a positive impact in some fields such as healthcare, but could also have broader societal impacts which may not all be beneficial.

• A good use to benefit mankind.

• There needs to be consensus about how AI is deployed

• Positive but it needs a fair amount of curbing/guidance and is vulnerable to learning from poor information as well as good info

• I do not know enough about it to comment.

• I feel quite unsure about it and apprehensive about any wide scale use without sufficient monitoring.

• I do not know enough about it.

• More positive than negative although I am aware that our private data is more at risk.

**Q3. Would accept HCP relying on AI to diagnose CD (n=67 comments)**

• Depends on how secure and accurate it us

• It's surely better than the enormous waste of my life that it took to get a diagnosis

• To rely on AI 100% could be dangerous

• As long as it was only used to inform the diagnosis - not the full diagnosis made with just AI

• Good training of healthcare professionals, backed up by appropriate AI would be better than total reliance on AI.

• Although a human opinion would be reassuring, using AI accurately would be more efficient, and accurate thus improve patient outcomes.

• In my case I couldn’t have a biopsy as I couldn’t deal with the camera so I had a pill cam.

• Don't know enough now.

• I would prefer a human looks first then run through AI

• I think so if it is really more accurate and faster.

• If properly tested through research

• After much testing and research

• AI Results should be reviewed by specialist.

• if I understood that the results are more accurate than a human alone

• Evidence to date indicates that AI can be trained to assess images and tissues far more quickly than a human and does not get tired or need a break. However, there is a need to monitor accuracy in case programming needs updating

• It is not safe to rely on anything in isolation. My mother nearly died because a doctor used a faulty thermometer on her, and paid heed to the result without taking account of the other signs that indicated there was a problem with the thermometer. Any user of AI needs not to use it blindly or without considering accompanying information.

• If it speeds up the diagnosis, then a good idea

• I believe this is the way forward to ensuring more people are accurately and quickly diagnosed as to whether or not they have coeliac disease. Either way, they can then make choices that will improve their health and/or have further testing and medical help. It should reduce waiting lists and waiting times.

• AI diagnosis would be a great start.

• I think AI could be a good guide but I would want them to 100% rely on it

• Only if I could be 100% confident that AI would produce a more accurate and faster diagnosis.

• If the success rate was higher than a human diagnosis

• If a trial showed accuracy in reporting from AI than yes.

• Needs more testing

• If the use of AI helps improve accuracy of diagnosis and in turn reduces waiting ss then it should be regarded as a positive step in lessening the impact of coeliac and other diseases.

• There has been little progress in diagnosis since being diagnosed with coeliac disease in 1990

• I would want to know that AI samples were regularly monitored by a human, to ensure its continued accuracy and to ensure the programme had not been corrupted

• As with a pathologist diagnosis I assume the option would still exist to seek a second opinion - probably at the patient's expense. How then would the definitive view rest between the AI and the pathologist? Possible shades of the Post Office's Horizon here if the base assumption is that the computer never lies. I am not sighted on how many edge cases there are where diagnosis could go either way - though I do not think that affects the fundamentals

• I went private for endoscopy to avoid having a 12 week wait and having to continue eating gluten when I was fairly certain it was CD

• If it sped up the process of diagnosis

• I think I would like a lot more information on how the AI determines diagnosis, as opposed to a pathologist.

• Proving the statistics demonstrate that the AI system is equal or better than the current system.

• So long as research can prove it’s more accurate/reliable

• Not as a solo reliance but if the trained healthcare professional concurred and this assisted them then why not!

• As long as the system has been robustly tested.

• I assume it would be backed up via blood tests

• I think it can be useful if the two work together. I don’t think AI should replace humans

• Would want more robust evidence that AI was able to do this conclusively

• However these system would have to be thoroughly validated

• When I was diagnosed my first biopsy said I did not have coeliac but as symptoms continued an endoscopy was performed and showed that I did. I am not sure if AI would show whether this was needed or just say the test was negative.

• But it should be backed up by other issues. i.e., if a GP suspects coeliac disease it could then be confirmed by AI.

• Early diagnosis of coeliac disease is key however would there be sufficient information for AI to accurately detect and diagnose?

• I'd say yes to "using artificial intelligence". If I was offered the use I would agree.

• As long as rigorous testing carried out to prove efficacy

• And the results could still be reviewed manually if there was any doubt.

• I can see AI playing a part but the determining mind should be the clinician, using all the observations and diagnostics and relying on their own knowledge and experience

• But backed up by human scrutiny

• There needs to be human QC built in to double check an appropriate proportion of AI diagnoses. Also suggest gradual changeover to AI to constantly review relative accuracy of both methods.

• Once the relevant AI software has reached an "adequate" level of quality and reliability.

• It depends on how accurate it would be.

• As mentioned I'd like confirmation by an expert

• After rigorous testing of its use

• Yes if there is a high degree of assurance that false negatives will be rare based on validation of the "software as a medical device".

• If my symptoms had been considered in the round over the years am sure it would have led to earlier diagnosis

• A little nervous

• I would be happy for AI to diagnose but would want a specialist to review and confirm diagnosis

• Provided that it has been properly tested and proven

• Definitely took until I was 41 before diagnosed.

• I feel that it would still be important for this to be overseen by healthcare professionals, even if at a system level.

• Better than uninformed professionals. In Bedford I have been diagnosed for years with IBS. When I told a cousin, a retired hospital matron, of my coeliac discovery, as I initially self assessed, she said had I still been in the West of Scotland, amongst the Irish/Scottish community, I would have been diagnosed years ago. Anecdote I know, but tolerance of industrially mass produced bread was not built up so quickly in the potato staple dependent community.

• Only if accuracy has been proven to be as good or better than a human pathologist

• As long as I was confident that there was research to support it and that checks were in place to monitor it

• It would depend on the accuracy of results gained over time

• I’m convinced I am coeliac positive even although my blood test has been negative

• After what I’ve just read it couldn’t be worse

• Nor reliant .. should use as support to speed up diagnosis

**Q4. Necessary for patient to be AWARE AI used (n=38 comments)**

• It is already being used in breast cancer imaging and matches or outperforms radiographers.

• Transparency is important. Also if symptoms persist and align with coeliac disease then option for a review.

• I have ticked yes as I feel this is generally good practice in relation to transparency by your medical professionals

• I'm sure that a lot of people are not aware that AI can help with e.g., cervical cytology screening.

• It would give me confidence that AI was being used to improve patient outcomes, improve accuracy, efficiency, effectiveness.

• Until it is widely used

• Some people might be sceptical that it could be a correct diagnosis

• We should be open and transparent about its use and the results

• Just in case there are issues that AI did not pick up. Human should always have the final say. Diagnostics is not as straightforward as might be thought

• I think we should all be able to know what tests are done on us, and how.

• Over time people will realise that there are some things that AI can do better than humans, leaving humans to do the more specialised and personal aspects of the job, like discussing it with the patient.

• I think that transparency is very important. Patients should always be provided with all information about their tests/treatment in order to help them make informed decisions.

• Some people simply won't trust or believe that AI is capable of making accurate diagnosis

• So many things are automated and we don't question them. People may only be unsure because it is relatively new.

• Assuming the accuracy of diagnosis remains high, then I don't believe patients need to be told. As an example, once a medication is approved to be used, patients are not told how the drug is effective, only that it works.

• The sooner one is diagnosed the better.

• If the AI has been thoroughly checked and tested prior to use, and it is regularly monitored, the AI system is part of the entire process and does not need to be mentioned. We all rely on machines daily and know they need monitoring/repairing/reconfiguring occasionally, but become an extension of our functioning capabilities.

• Arguments both ways here but thinking about the current system, to the patient it is a black box. An AI based diagnosis is still a black box. The principles of diagnosis are presumably the same in both cases. So why does it fundamentally matter? A caveat would be how to maintain the independence of the AI model. If say the NHS wanted to keep the number of diagnoses low? Arguably that scenario could arise now.

• Together with the AI versus human detection %

• Yes and exactly what role it was playing in diagnosis.

• I’m certain that being diagnosed is primary factor

• Patient should be able to access the pathology report, and this should make it clear that the report is AI generated (in lieu of having the reporting pathologist's name as per usual)

• Any medical condition diagnosed (or test results coming from) AI should be communicated to the patient in my opinion.

• It is probably desirable but that knowledge needs to be put within the context of the accuracy of conventional diagnoses

• It is important that the individuals are informed when AI is used for medical purposes.

• Some patients might object so I guess they should be informed

• I believe patients need to be always informed about the tools leading to their diagnosis.

• Patients should always be made aware of, and agree to any and all matters referring to their health

• To allow for choice and/or as a 'second' opinion, however I expect trials will determine which is the most accurate process for testing?

• Hiding this information would be paternalistic on parts of the NHS, but on the other hand the scepticism of some people towards these technologies may lead to them not accepting the diagnosis.

• Always inform

• Yes, at least in the initial stages of adoption of this new technology.

• Personally I feel it's more important as a patient to understand the likelihood of misdiagnosis - especially false negatives.

• For me I'm not fussed

• Yes, only because human checks need to be made from time to time.

• Is a question for consideration, whether a professional recognises the need to seek AI support. Of course I do not know, and my experiences of GP's is they do not recognise coeliac as the potential problem.

• I'd prefer to be aware but I'm unsure what other people might think. I'd like to know the proven reliability of the AI system, though I've never questioned the reliability of pathology staff. Interesting question

• I think it is always important for a patient to fully understand their care, including the process of diagnosis.

**Q5. Necessary for patient PERMISSION for AI use (n=34 comments)**

• as new and unfamiliar technology - over time this would fall away

• I would not think permission is required but as indicated in response to Q4, openness and transparency it to be welcomes

• Would like to be informed of AI being used in diagnosis

• If the AI has been tried and tested as an aid to well trained healthcare professionals it might speed diagnosis.

• AI should be part of medicine, to improve Patient Outcomes, efficiency, effectiveness.

• Need all the information that is available.

• I assume any tests done are completed in the most professional manner

• Depends on the patient

• With the best will in the world most patients will not know enough about AI to make a decision (though they may THINK they do!)

• I do not see how this would help a patient.

• If you have the evidence to show that the method you are using is the best one, then that is the most important thing. I do not expect to state what type of test is to done when looking for indicators of a specific illness - I expect the qualified expert to know what to do.

• The NHS is a free service and people need to accept that what is cost-effective is what should be used as long as the results are at least as good as previously used methods.

• After all they can only say No or Yes

• No (see answer to Q4.)

• Same as above

• As long as IT systems are secure

• See comment on 5.

• Maybe during the inception as we get used to the idea of AI being used in this manner. However I am very pro the use of AI as more accurate than humans.

• Patients aren't asked to consent to which pathologist will review the sample, or which tools may be used in the slide prep, or which microscope will be used to view the sample. AI in this setting sits somewhere between being a pathologist and a tool of the lab, and patients don't explicitly give consent for either of those things currently, even if it is a new pathologist or a new brand of microscope. It is therefore not logically consistent to expect consent for a new AI model.

• I think if the AI is better at diagnosing health conditions, they should be informed that AI will be used in the testing process.

• Not if the trained professional is making the formal decision based on the AI work.

• The pros and cons should be explained and a right for a second opinion if requested

• Anything out of the ordinary should require permission. A biopsy anyway should require permission.

• But need to be informed if the diagnosis system is well set and validated. Permission should be obtained if for research purposes

• With explanation

• I think if a patient was asked for permission some people might not agree to it. I personally would give permission but some people are still unsure about AI.

• As long as the diagnosis is then confirmed by a pathologist, then there should be no issues.

• Certainly in the short term until AI is shown to be unequivocally at least as accurate as human review.

• If this new technology proves reliable, then there should be no need for permission. I guess all sorts of modern medical equipment is used without prior patient's permission.

• If the system is approved and validated for intended use.

• Wouldn't bother me

• Yes

• It would be polite to let them know and explain the benefits

• Not needed if AI is well proven to be as good or better than human pathologist

**Q6. Trust in AI diagnosis if approved by regulators (n=48 comments)**

• How safe and secure would it be

• Providing i could see written results to consider them myself

• I would have more trust in artificial intelligence system - humans missed my symptoms for 40 years!

• In my diagnosis the biopsy was inconclusive but the blood tests were positive both times. there will always be uncertainty in any system - i decided that the blood test was enough to accept the diagnosis

• Define who the necessary bodies are, please!

• This could be a more reliable method than humans who can disagree with findings.

• It took decades for me to be diagnosed due to human error throughout, and I have been very disabled, ill with Gluten Associated Disorders including Neurological damage, Cerebellar Ataxia, Osteoporosis, it has had a dreadful effect on my body and life, AI hopefully would stop the medical human error, and the medical bias into wrongly believing everyone has a functional disorder rather than a autoimmune disorder than can be treated with the right diet to prevent harm, disability. AI would improve Patient Outcomes, that must be the goal.

• Again I do not know enough about AI but am willing to keep an open mind until I understand more about it.

• If all proper research completed

• You are the experts!

• It seems to me that AI should be used in conjunction with other diagnostic tests i.e. health/symptom history and blood tests

• I would be happy with this

• I would need to know details of the quality of the research, the numbers it had been done on, whether those who had been tested represented me (age, sex, ethnicity, BMI, comorbidities etc.

• As long as the regulatory body is interested in quality of results and not just cost-savings

• As above. Only once fully tried and tested.

• I’ve had countless blood tests and 2 gastroscopies to diagnose coeliac disease but all negative. I now know that I do not have the gene for coeliac disease but there is a strong possibility that I have NCGS (no test as far as I am aware). Symptoms all gone after 6 months gluten-free diet.

• I would say very strong, but AI can have failures too

• Same as above

• The criteria for approval would need to be consistent with generally accepted medical practice.

• Based on %

• I would want to know the research results on which its use was being based. If it’s 95% accurate vs 85% for a pathologist (tho I’m confused about how the control/baseline worked in those figures in relation to this project) then I’d be reasonably happy about it.

• As long as it has been proven

• If it's passed regulatory approval then at a minimum it is non-inferior, if not superior to pathologist...

• My GP overlooked a marker in my blood test in 2014. I ended up getting diagnosed in 2020 after years of suffering with symptoms so humans can make errors in the same way AI might

• A diagnosis by AI is fact based and not subjective.

• Providing the patient is fully informed and there are quality checks by an expert

• I would like human input as well in the diagnosis process

• Would need to be convinced that there was no room for error in the diagnosis

• Because a regulatory approve AI application for clinical use will still require validation of sample with less predictability

• I see comments on AI solely diagnosing coeliac disease

• a lot of research is put into place before any form of system is put into place and the results from this research show it works.

• I don’t see it as being particularly different from the trust we place in other accredited automated tests.

• As i say above, I can see AI playing a part but the determining mind should be the clinician, using all the observations and diagnostics and relying on their own knowledge and experience

• If proven to be better than human intervention.

• Strong, as long as the regulatory body sets out very stringent approval criteria.

• After an appropriate level of checking the AI results again the current process

• Again, it would be how accurate the results would be. And is there any checks made by a person too

• Like all AI it still needs testing at this stage

• Am sure there would be extensive trials prior

• A computer system doesn't get tired or distracted

• My concern is not the use of AI as such but the competency of the regulatory bodies to assess an AI system and ensure it has been tested and proven.

• As long as regulatory bodies are robust

• The level of confidence in a diagnosis made using an AI system would clearly depend on its performance levels, so this response assumes that it would perform at least as well as a medical professional.

• It can enhance the current situation.

• I do not know enough about AI. I am 66 and a bit “traditional” and not tech-minded. However I can see that this is the way forward.

• The longer a track record it has, the more I'd trust it

• Maybe it would give a diagnosis quicker than the doctor

• If 95% accuracy can be maintained.

**Q1. Prior awareness of AI (n=28 comments)**

• Can’t remember

• Academic study

• Combination of news articles i have read etc

• Through workplace

• I work in AI for medical research

• Local Health Board contacted me for my views about this.

• Social media

• Christmas lectures on tv

• Work for NIHR

• Intelligence of Software, machines etc

• through my field of work

• So long ago I can't remember!

• Not sure - been aware for very long time

• Just grew up always knowing it existed

• Work

• Through work

• News on TV

• Work

• General understanding of developments in IT and work on decision systems during my career

• Various places, books, movies, news

• I am a researcher in AI

• University education

• Radio 4/ World Service

• News and work environment (software & hardware).

• IT background

• Have done some work with it

• Teach Adobe software with some features that use a very limited form of AI

• Friend’s son was developing years ago

#### Supplementary Table 3

**Associations between demographics and questionnaire responses - national online survey (n=231)**

Exploratory analysis of associations between three demographic variables (age, gender, highest level of education) and responses to the six questionnaire items (Q1–Q6; see Supplementary Table 1 for full response distributions). For each item, the primary omnibus test is shown for each demographic: Kruskal-Wallis for age and education (both treated as ordinal/continuous), and chi-square test of independence for gender. Spearman rank correlations were additionally used for the two fully ordinal items (Q2, Q6), shown in the final two rows. p-values are corrected for multiple comparisons across all 22 tests conducted (the 18 primary omnibus tests plus the four secondary Spearman trend tests) using the Benjamini-Hochberg false discovery rate procedure; bold cells denote associations that survived correction (p-adjusted < 0.05).

| **Questionnaire item** | **Age (Kruskal-Wallis)** | **Gender (Chi-square)** | **Education (Kruskal-Wallis)** | **Direction of statistically significant effect** |
| --- | --- | --- | --- | --- |
| Q1. How first heard about AI | H(5)=11.64 p=0.040 (adj. 0.102) | χ²(5)=12.10 p=0.032 (adj. 0.100) | **H(5)=23.53 p=<0.001 (adj. 0.006)** | **Education:** More educated respondents more often cited a scientific publication or forum (18–20% of postgraduate/doctorate holders vs. 0–3% of those with school-level education only) and less often reported never having heard of AI before (0–4% vs. 12–22%). Age (p=0.040) and gender (p=0.032) also showed raw associations with Q1 that did not survive correction (adj. p=0.102 and 0.100 respectively). |
| Q2. Feelings about AI in daily life | H(4)=7.57 p=0.109 (adj. 0.159) | χ²(4)=6.06 p=0.186 (adj. 0.228) | H(4)=10.67 p=0.031 (adj. 0.100) | Not applicable – education showed a raw association on both the omnibus test (p=0.031) and the Spearman trend (p=0.042, see final rows) but neither survived correction (adj. p=0.100 and 0.102 respectively). |
| Q3. Accept clinician relying on AI | H(2)=1.83 p=0.400 (adj. 0.417) | χ²(2)=7.25 p=0.026 (adj. 0.100) | H(2)=5.35 p=0.069 (adj. 0.147) | Not applicable – gender showed a raw association (p=0.026) that did not survive correction (adj. p=0.100). |
| Q4. Awareness of AI use necessary | H(2)=9.26 p=0.010 (adj. 0.054) | χ²(2)=5.11 p=0.078 (adj. 0.147) | H(2)=3.07 p=0.216 (adj. 0.250) | Not applicable – the raw association with age (p=0.010) does not survive correction for multiple testing (adj. p=0.054). |
| Q5. Permission for AI use necessary | H(2)=3.59 p=0.166 (adj. 0.214) | χ²(2)=2.89 p=0.235 (adj. 0.259) | **H(2)=11.62 p=0.003 (adj. 0.022)** | **Education:** Perceived necessity of patient permission fell with education: 77.8% answered "yes" among those with no formal education, vs. 25.0% among doctorate holders. |
| Q6. Trust in regulator-approved AI | H(6)=11.00 p=0.088 (adj. 0.147) | χ²(6)=5.92 p=0.417 (adj. 0.417) | H(6)=9.69 p=0.138 (adj. 0.190) | Not applicable (see secondary trend test; final row) |
| Q2 (secondary trend test). Feelings about AI in daily life | Spearman ρ=−0.11 p=0.089 (adj. 0.147) | - | Spearman ρ=0.13 p=0.042 (adj. 0.102) | Not applicable |
| Q6 (secondary trend test). Trust in regulator-approved AI | Spearman ρ=−0.11 p=0.094 (adj. 0.147) | - | **Spearman ρ=0.20 p=0.003 (adj. 0.022)** | **Education:** Trust in AI-assisted diagnosis increased with education level (e.g. "very strong" trust reported by 11.1% of those with no formal education vs. 37.5% of those with a doctorate). |

**Notes**

1. Establishing direction of effect that survived correction for multiple testing: The omnibus tests above indicate whether an association exists but not its direction. For the three associations that survived Benjamini-Hochberg correction (Q1×education, Q5×education, Q6×education), direction is evident directly from response percentages across education's ordered categories, shown above and in the final row. Q4×age was significant on the underlying Kruskal-Wallis test (H(2)=9.26, p=0.010) but did not survive correction for multiple testing (adjusted p=0.054) and is therefore not reported as a significant finding.
2. Effect sizes for the two Kruskal-Wallis associations that survived correction (Q1, Q5) were small (ε²=H/(n−1)=0.05–0.10); the Spearman trend for Q6×education was ρ=0.20. These associations, while statistically robust, should not be read as strong predictors of individual attitudes.
3. Four gender chi-square tests (Q1, Q2, Q3, Q6) had an expected cell count below 5 in at least one cell, which can make the standard chi-square approximation unreliable. A 5,000-iteration permutation test was run for each as a validity check and closely matched the asymptotic p-value in every case (Q1: 0.033 vs. 0.029; Q2: 0.194 vs. 0.185; Q3: 0.027 vs. 0.026; Q6: 0.432 vs. 0.413; asymptotic vs. permutation), indicating this caveat does not change any conclusion.

#### Supplementary Table 4

**Stevenage focus group (14 March 2024) - raw discussion notes as recorded by facilitators (anonymised)**

**Getting a diagnosis - negative:**

• Long time to get a diagnosis from onset of symptoms.

• “Looking back I had symptoms for maybe 20 years prior to my diagnosis”

• Time from noticeably feeling unwell to getting a diagnosis varied greatly between individuals.

• Variations in the diagnosis pathway; in one focus group six were diagnosed with blood test followed by endoscopy, two with two blood tests and one with just one blood test.

• “I was placed on a gluten free diet during Covid and then had to participate in a gluten challenge and re-introduce gluten post Covid to achieve a medical diagnosis”

• Long wait after blood test to get an endoscopy to complete a diagnosis.

• GPs often could not identify the problem at first, initially dismissing many of the individuals. Most people reported needing to see their GP multiple times before coeliac disease was considered or tested for.

• For some, diagnosis was only eventually given when they changed GP because of moving house or a new GP joining the practice.

• “It was only when I moved back to the UK under a new GP that I was tested (for coeliac disease)”

• “I had a blood test that was positive but no one said anything until I was really poorly some years later and then I was invited for an endoscopy”

• “I was diagnosed a year ago. I had anaemia, was losing weight and had rashes. It took four months to get my blood test results and another 9 months before I had my endoscopy!”

• “My brother was diagnosed and as a family member I was screened, but I didn't have my endoscopy and biopsy until several months later. It then took 3-4 weeks to get the results. The whole process took a really long time but at least I squeezed in another Christmas before having to go gluten free!”

• “I found it hard to accept the diagnosis as although I have family members diagnosed with the condition I have no symptoms, I couldn't interpret the results in the letter I received and having a visual result would have been helpful. I doubted the diagnosis but realised this was only wishful thinking!”

***Often misdiagnosed (e.g., IBS, diverticulitis)***

• “I was diagnosed with diverticulitis over the phone after experiencing troubling gastrointestinal symptoms for about 1 year. Sometime later, someone from my GP practice phoned me up asking me to have a blood test to rule out other symptoms. That blood test showed I had coeliac disease, and I was referred for an endoscopy to confirm this.”

• Many chose to have a private biopsy due to long waiting times on the NHS.

• “After a positive blood test, I was referred for endoscopy and told it could be a few months before I would be seen so I chose to go private.”

• Parents had to be resilient to get a diagnosis.

• “I noticed he had stopped growing and had very bad brain fog, with barely any memory and he was struggling at school. I was repeatedly told he was ‘just slower than other kids’ and it was ‘nothing to worry about’!”

• “I was told my daughter was just growing more slowly than her cousins of a similar age”

**Feelings at diagnosis - positive**

• Relieved to finally have an answer, be taken seriously. Not 'in my head'.

• Relieved that it wasn't something more serious e.g., cancer.

• Relieved that there was a way to begin to feel better and alleviate symptoms.

• Accepting of diagnosis, as symptoms aligned with what they had been experiencing and symptoms eased when following a gluten free diet.

• “Within a couple of months on a gluten free diet (my son) was growing rapidly and the brain fog lifted. Now whenever he's glutened he can barely remember his own name”

• “(my daughter) made remarkable improvements on a gluten free diet”

• “I looked the symptoms up online and they matched how I'd been feeling”

• “If I hadn't felt better following a gluten free diet I might have questioned the diagnosis”

• People who had 2 x blood tests and an endoscopy felt assured that the diagnosis was correct.

• “I was shown the captured images of my gut after the endoscopy and they explained the damage so it was helpful for me in accepting the diagnosis.”

***Feelings at diagnosis - negative***

• If diagnosis seemed rushed or hadn't followed best practice guidance, less likely to accept and felt doubtful that the diagnosis was true.

• “I only had one blood test and nobody explained what they were testing for or gave me my results so I didn't know for almost a year. I only found out because my GP practice called to offer me the flu jab and I asked why I was eligible as I didn't normally have it and she told me my last blood test had indicated coeliac disease. I'm not following a strict gluten free diet”

• Worried for family members, as there is a genetic link.

• “I received a letter that included images of my gut and the antibody levels but there was no explanation of the results just that they were positive”

• “I was in my mid-twenties when I received my diagnosis, I was really upset but learnt to accept it.”

**AI (general awareness)**

• Majority had heard of AI.

• Many were aware of the application of AI in modern society and had positive things to say e.g., phones.

• Some recognised its introduction into healthcare via the news.

• Some noted that it was viewed quite negatively.

• “it gets bad press”

• One person mentioned that they had a heart condition and the nurse had mentioned to her that her scans were to be assessed with AI.

**AI in coeliac disease diagnosis - negative**

• Concerns about lack of human oversight.

• “I would want a human to check my sample”

• “I would doubt an AI diagnosis more quickly than a human diagnosis if I wasn't beginning to feel better following a gluten free diet”

• Diagnostic delay is due to the waiting times to have an endoscopy and biopsy, not to receive biopsy results - is the purpose to free up time and reduce the waiting time to get a diagnosis or to increase accuracy of diagnosis? It's both.

• “They told me straight after my endoscopy that it was clear I had coeliac disease, but they would send the biopsy off anyway and I would receive a call to confirm. I got the call 3 days later”

• “I got the results of my biopsy 3 hours after leaving the hospital”

• “it would be better to invest time and money into training pathologists better so there is less variation”

***AI in coeliac disease diagnosis - positive***

• Agree it could eliminate the subjectiveness and variation of diagnosis by pathologists.

• “Why would you choose a less accurate diagnosis (human) if there are more accurate options available (AI)?”

• “Machines are objective, humans are subjective”

• Many agreed they would accept a diagnosis by AI if it was approved by a rigorous standard and appropriately certified, with acceptable quality assurance/control methods in place, and active reviewing of parameters. Would accept if started to feel better on a gluten free diet.

• It could be cost-effective for the NHS and free up valuable human time.

**Consent, ethical and other considerations**

• Majority would want to be told if their diagnosis was done by AI.

• Some information would be useful about what Marsh scores mean and what their biopsy score was, e.g., some blood tests give you your results in relation to the parameters, or you're shown your ECG graph.

• “Some information is useful for example blood test parameters but you don't ask how the test was done”

• AI to only be used if pathologists disagreed.

• “You don't normally ask your doctor to explain how the diagnosis was made”

• Majority would trust/accept diagnosis by machine alone if more accurate and certified appropriately.

• “We trust lots of health tests which use machines and tests that aren't 100% accurate like blood tests even”

• “Nothing is 100% accurate but we still accept diagnoses”

• Some mentioned that it would be good to have some info on how the machine/system works. Others argued it is unnecessary.

• “You don't ask to speak to a pathologist to ask them how they came to that decision”

• Some were concerned that if there were differences in pathologists' decisions on diagnosis, how could they be reassured that the information used for programming the AI solution is accurate in the first place.

• There was a desire to know what other information would be used alongside the AI for assessing biopsy samples, e.g., would the programme have information about whether there was a family history, the antibody test results, to further assist in the diagnosis?

• There was also a desire by some to know what checks and balances would be in place, as there was an expectation that the AI system would be further developed the more it is used. How would updates to the software be issued and how would this be controlled across different testing centres to ensure they are using the latest version? Some felt there was the same risk in humans, as some rely on what they were taught in medical school, whereas others are continually reading up on research and making sure they are up to date with the latest best practice.

• Some wanted to know when human intervention would take place, e.g., would there be a regular scheduled QC check by a human, would the AI system flag grey-area cases for further assessment by a human.

• Many wanted reassurance that other conditions wouldn't be missed if this AI diagnostic pathway was adopted. They also wanted to know how the results would be shared with HCPs and patients, considering there had been many different experiences to date, most of which had been confusing, and they wanted this to be avoided in the future.

• Everyone recognised that the wait time is really associated with pre-endoscopy and biopsy, so the key selling point for the AI technology is the potential improved accuracy and release of pathologists' time and therefore potential cost savings.

**Thoughts about healthcare professionals**

• Many felt that it would be important to have GPs and other healthcare professionals who are involved in the diagnosis pathway for coeliac disease informed early on in development, so that they could get onboard from the start.

**Individual diagnostic journeys (facilitator notes, redacted for identifying detail)**

• One male was feeling lethargic and lost weight, didn't feel like himself for around 2 years prior to diagnosis. Visited the GP but the GP did not know what was wrong. As symptoms didn't improve and time went on, they began to assume the worst e.g., cancer, and began to fight harder to be listened to by their HCP. Eventually the GP decided to request a blood test for CD. This came back positive, so the patient was referred to see a gastroenterologist for an endoscopy and biopsy. The patient cannot recall how long they were expected to wait exactly but remembers it was a few months, so they chose to go private to be seen quicker. Once diagnosis was confirmed privately, they were referred back to the NHS. As his symptoms matched his diagnosis, he accepted it. He was relieved it was not something life-threatening and that it was something he could adapt to manage. When symptoms improved quickly after beginning a gluten-free diet, it solidified his acceptance of the diagnosis.

• Another male patient experienced similar symptoms of lethargy and began to lose weight some decades ago. He was dismissed repeatedly by the GP until someone thought to do a blood test for CD. This was positive and he was referred to a gastroenterologist for an endoscopy and biopsy which confirmed the diagnosis. As the blood test and biopsy were done and his symptoms matched those of CD, he accepted it. If he hadn't felt better on a gluten-free diet he may have challenged the diagnosis a few months later. The patient reports that he was given gluten-containing pasta as part of a meal post-operation in hospital, despite being there for suspected CD.

• Majority of attendees reported having to visit the GP multiple times as they were initially dismissed and not listened to. It took more than one visit for the primary care practitioner to consider CD, for both adults and children.

• One person reported that they never doubted their diagnosis, as they were told the symptoms at point of diagnosis and they matched what they had been experiencing, so it seemed plausible. They later looked up the symptoms online to further confirm that CD aligned with the symptoms they had been experiencing.

• One male was originally misdiagnosed over the telephone, during a period of lockdown, by his GP with diverticulitis after experiencing symptoms for about 1 year. Some years later, someone at his local practice noticed that he had not had a blood test to rule out other conditions before being given the diagnosis, so a blood test was done to test for CD which came back positive, and he was referred to a gastroenterologist for an endoscopy and biopsy. He chose to go private to receive a quicker diagnosis. As he began to feel much better on a gluten-free diet he knew that the first diagnosis was wrong and accepted a diagnosis of CD.

• Another male reported that it took around 20 years for him to receive a diagnosis, as he originally approached his GP with symptoms but was dismissed as having IBS. He then suffered with this whilst living abroad, and only once living back in the UK and complaining to his new GP about his symptoms was he tested for CD by blood test. Once beginning a gluten-free diet he felt a lot better and therefore knew the diagnosis was correct.

• The majority of people felt relief once diagnosed - that it wasn't something more serious and that they had an answer, it wasn't 'in their head' and that their symptoms could potentially be controlled. The majority reported immediate acceptance of their diagnosis, which was solidified by the fact that they began to feel better when eating gluten-free.

• One female was in doubt about her diagnosis as she only had 1 blood test and was not formally told her diagnosis, nor given any support to begin a gluten-free diet. She went to the GP with various symptoms, so the GP ordered a blood test but didn't tell her what would be tested. Some months later, she was invited for the flu jab and when she questioned why, as she was young and was not normally offered it, she was told over the phone that she has CD. As she only had the one blood test and has not been offered any support, she does not fully accept this and has not begun a strict gluten-free diet to see if this eliminates her symptoms.

• One mother was concerned about her baby, who had seemed poorly almost since birth, always being sick and having a very bloated tummy. Eventually the GP tested her for CD and this was confirmed by endoscopy. She never doubted her daughter's diagnosis and began a gluten-free diet immediately, after which her daughter made remarkable improvements, so they knew it was correct.

• Another mother noticed her son was not growing very quickly, at a young age. He was also very lethargic and experienced very bad brain fog, having barely any memory, struggling terribly at school. She was constantly at the doctors trying to find out why this might be and had to really push to be listened to, but was told he was just slower than other kids, nothing to be worried about. Eventually, a couple of years later, a new GP tested him for CD via blood test, which was positive, and he was diagnosed with two blood tests and began a gluten-free diet at around age 10. She didn't doubt the diagnosis and was just relieved that they had an answer. Within a couple of months on a gluten-free diet he was growing rapidly and the brain fog lifted. Her father had CD but as he didn't have the same (typical gastro) symptoms she didn't suspect it could be that, so never pushed for him to be tested for that. Luckily the new GP asked about family members and made the connection. Now, when he gets glutened, he has terrible brain fog and 'can barely remember his own name', so they have no doubts as to his diagnosis. He receives good follow-up care.

• One lady was told that her daughter was just smaller than other kids, as she was much smaller than her cousins who were born around the same time, and had to push to be listened to.

***Breakout discussion: AI in the diagnosis of CD (second note-taker)***

• One individual stated that AI should be used in diagnosis if it's more accurate than the current gold standard of diagnosis. Why should we choose to be diagnosed by a less accurate method if there is a better, well-tested option available? Others in the group indicated agreement with this point - accuracy is an important consideration.

• Some expressed concern about the lack of human control and oversight.

• One person stated that as the blood test is considered accurate enough to diagnose some people, including children, without an endoscopy and biopsy, research should be focusing on using a blood test alone for the diagnosis of everyone.

• The risks associated with using AI should be identified and accounted for and mitigated/managed appropriately.

• The benefits should significantly outweigh the risks of the current gold standard.

• Someone proposed the idea that humans should check what the AI system suggests to ensure it is true. Someone else said that this would be duplicating work; there would need to be a balance, as it would reassure people if there was some human oversight, e.g., every 30 slides, a human pathologist checks the decision to see if they agree.

• Someone proposed the idea that it may be more worthwhile to invest time, money and resources into understanding the variance between pathologists and to work on better educating and improving this gap. Pathologists should be keen to be shown their mistakes and to understand why they got a diagnosis wrong so they can look to improve and this variance can be reduced.

• Someone said that people might initially accept a diagnosis by an AI system but be more doubtful, more quickly, than if they were diagnosed by a human pathologist. For example, if their symptoms didn't reduce very quickly on a gluten-free diet, they might return to their GP and think the diagnosis is wrong, when if they were diagnosed by a human they might wait longer to question it. This can be resolved by education at diagnosis, e.g., how many months for symptoms to reduce.

• Many people agreed that, like with a human diagnosis, if you start to feel better and symptoms match the diagnosis, it will be accepted.

• One individual felt that research should be focused on improving aftercare and ensuring patients who are already diagnosed get proper follow-up care and are not forgotten about.

• One person said we could be using AI to ensure people get the best support post-diagnosis.

• One person stated that they are unsure how it would help people get a diagnosis quickly, as it is more the wait to see a gastroenterologist for the endoscopy and biopsy that is the issue, not the wait time for a biopsy to be viewed by a pathologist.

• Many of the patients commented that their diagnosis had come very quickly after the endoscopy and biopsy, with the maximum reported being 2 weeks and most reporting a couple of days up to a week. A few commented that the gastroenterologist had told them their diagnosis and to begin a gluten-free diet at the same appointment, but that they were sending the biopsy off anyway.

• One person commented that the AI system should be used to identify the uncertain cases - the cases where humans would disagree or be unsure.

• Machines are objective and humans are subjective, so the machine algorithm is very beneficial.

• One person commented that they would want a bit of information about the machine.

• All agreed that they would want to know that it was done by a computer system and not a human pathologist.

• One elderly gentleman commented that he was okay with a machine looking at his biopsy and suggesting a diagnosis, but that he would then want this checked by a human before he would accept the diagnosis and commit to a gluten-free diet.

• Someone commented that when you receive a diagnosis done by a human, you accept it. You don't normally ask to speak to a pathologist or other HCP, as you know that they and the tests they use are adhering to a rigorous system.

• When you have an ECG it's comforting to be shown the graph as the doctor explains it and can point out things. It would be helpful to have a paper report, or maybe an image of the slide that the AI system looked at, shown to you, to help you accept the diagnosis when it is given to you.

• When you get a blood test you're shown parameters, e.g., this is the normal range. Maybe a similar approach could be adopted, e.g., this is the image, the system stated Marsh score X, showed villous atrophy, etc., which means you have CD.

• Someone said they would trust it. They wouldn't expect it to be 100% accurate either, as nothing is. We trust hundreds of other tests relating to our health, e.g., blood tests, which aren't 100% accurate as that is impossible to achieve. It should have a very high level of accuracy, above that of a human, and under proper clinical validation to be acceptable.

**Structured discussion notes (second facilitator, by topic)**

***How long did it take for diagnosis?***

• Biopsy - years to get any questions answered - there was no blood test.

• Diagnosed by symptoms only - they couldn't do an endoscopy.

• 5 weeks.

• Picked up by anaemia through the GP.

• Grew out of it as a child and then the GP put him back on gluten.

• Had a blood test for something else and picked up with a diagnosis some months later.

• Endoscopy at an NHS hospital.

• Diagnosed with lactose intolerance some years prior; blood test and endoscopy followed some months apart. Presented with lots of symptoms but not added together.

• On iron tablets for years on and off. GP suggested coeliac disease.

• Daughter had a blood test; the appointment took 6 months and then the results were not complete so the diagnosis was questioned.

***How did you feel on diagnosis?***

• Relieved that it wasn't cancer.

• GP told him he didn't have CD, and then consulted a colleague and rang him back and told him otherwise.

***Everyday problems***

• Expense of diet.

• Didn't bother with a nutritionist, but she did call the patient to check she was coping.

• Addicted to reading labels.

• Different health authorities - so may live in one but receive treatment from another.

• Made her more interested in gene and autoimmune research.

• Would love a discount card for supermarkets.

• Always offered vegetarian - places need more catering guidance and offerings.

***What did they know about AI?***

• All had heard of AI.

• Those who were in education were familiar with it for classroom and teaching use and writing reports and letters.

• All smartphones use AI but people don't question it.

• Apps.

• Use in cancer treatments - interestingly no-one mentioned diabetes or any other autoimmune condition.

***AI in the future***

• All felt it would be useful for repetitive tasks and mathematics but there is a fear of complete human replacement.

• Counting cells - AI clearly good for accuracy and repetition and removes the human difference of opinion.

• AI good for 80% to 95% accuracy but is it to free up time or to increase accuracy?

• They were shocked that analysis by pathologists takes only about 5-8 minutes per sample and that only 5% of pathology departments were fully staffed.

• Majority said that they would want to be told if they were using AI for their samples, but they agreed that it was generational - the older members would prefer a human, but all felt that the process needs to be explained.

• Concern about possibility of hacking the software and a QA query.

• They understood about the testing and that it was similar to trialling a new drug - real-life testing to improve condition/prevention/repetition.

• Improving by redoing the process.

• Certification.

• Active reviewing of parameters.

• Doctors often go by probability whereas AI is more mathematical.

• Gene therapy question - why do some children have it and some do not. Conditions change over time as children get older.

• Most would be accepting of AI as a second opinion but would also accept it more than a GP.

• Accepted that they still needed a human to analyse and deliver results.

• AI is a tool.

• Medical professional needs to be able to interpret, so training is paramount.

• AI would reduce the length of time of diagnosis.

• It would be cost-effective for the GP and NHS.

• Frees up medical staff as researchers use the AI.

• Most understood that CD diagnoses were pushed back if a patient presented with cancer, for example, but using AI would mean that cases could be prioritised and screened.

#### Supplementary Table 5

**Bury St Edmunds workshop (14 June 2025) - thematic answer summary (20 completed questionnaires)**

**Why does it take so long to diagnose coeliac disease?**

• Symptoms non-specific/vague; gradual symptom onset; variation between individuals

• GPs not always educated about symptoms/risk (incl. family history); CD not considered as explanation for symptoms

• Diagnosis imprecise, often not definitive, and invasive; difficulties with biopsy interpretation

• Lack of screening and funding; NHS waiting lists cause delays to see a specialist

• Patient-side factors: denial, fear, not wanting to ‘make a fuss’, reluctance to eat gluten before biopsy, non-compliance with diet

• Being labelled as IBS / dismissed (“it's your age…”); lack of community awareness

• Misdiagnosis of dermatitis herpetiformis; lack of joined-up care

**Who needs convincing that the method works?**

• Patients, relatives and the general public; pathologists and other clinicians; AI sceptics

• Government (Dept of Health/Education), regulators, NICE, NHS commissioners

• Funders: charities, companies, philanthropists, research committees

**How might we convince them?**

• Media publicity and public talks; social media awareness campaigns

• Patient testimonials on quality-of-life improvement; philanthropist/personality endorsement

• Demonstrating test performance (sensitivity/specificity) on a large confirmed-case series

• Lobbying MPs and involving Coeliac UK; framing around reduced long-term NHS costs

• Reassurance that a human remains in the loop

**Perceived risks (‘potholes’) and suggested improvements**

• Funding and commercial viability; competition from other groups

• Training needs (more pathologists trained to use AI); pathologist/GP resistance over job security

• Trust: risk that a poorly performing early AI tool could damage broader confidence in AI

• Regulatory approval; risk of interference reducing AI accuracy; need for the work to be checked

• Resistance to change, inertia, lack of enthusiasm from non-specialist funders

• Suggestion: screen family members given hereditary risk

#### Supplementary Table 6

**Edinburgh focus group (26 September 2025) - raw discussion notes as recorded by facilitators (33 attendees; 27 responses analysed)**

**In your view(s), why does it take so long to diagnose coeliac disease?**

• Lack of awareness (public and medical)

• Vague / non-specific symptoms

• Misconception that it is a childhood disease

• No 'classic' stomach symptoms

• Non-classic presentations (migraine, dermatitis, anaemia)

• Misdiagnosis as allergies / histamine intolerance

• Reliance on GP's personal knowledge of coeliac

• GP refusal to test without classic symptoms or strong family history

• Symptoms similar to other conditions (IBS, IBD, Crohn's, colitis, endometriosis)

• Not being listened to

• Unrecognised symptoms

• Symptoms normalised (stress, ageing, menopause, anxiety)

• Gender bias (anaemia in women attributed to periods)

• Reluctance to see GP

• Difficulty booking appointments

• Inability to see same GP consistently

• Overworked GPs

• 'Treat first, diagnose later' approach

• Symptoms must be severe before testing considered

• Misdiagnosis and incorrect treatment

• Treating symptoms not root cause

• Lack of frontline staff knowledge

• Too much research based on men

• Connection with other autoimmune disease not made (e.g., Graves, thyroid disease)

• Coeliac only considered after other diagnoses

• Low blood results but very symptomatic

• Symptoms vary greatly / inconsistent

• 1/3 asymptomatic

• Mild or transient symptoms

• Symptoms erupting post-pregnancy

• Symptoms overlapping with other conditions

• Negative or borderline TTG results

• False negatives

• Genetic (HLA) testing not offered

• EMA positive but not acted upon

• Inconclusive results delaying diagnosis

• Variation in testing protocol (doctor to doctor / NHS trust to trust)

• Blunt instrument of TTG testing

• Blood tests do not routinely include coeliac

• Multiple negative tests causing delay

• Tests negative before later positive

• Not told tests are for coeliac

• Not told tests may need repeating

• Not understanding need to eat gluten before testing

• Gluten-free diet started before testing

• Reluctance to reintroduce gluten for biopsy

• Amount of gluten required before testing unclear

• Wait times for blood tests

• Time to pathology read-out

• Wait for endoscopy

• Surgical / endoscopy waiting lists

• NHS cuts / NHS overloaded

• Delay between test and diagnosis (e.g., several months)

• GP processes causing delay

• Diagnosis quick once biopsy completed

• GP suspecting bowel cancer before coeliac

• Anaemia attributed to diet (eat more red meat)

• Diagnosis pushed forward by patient / friend support

• Lack of joined-up care

• Differences in follow-up practices

**Who do we need to convince that our method works?**

• Whoever commissions the test (GP / clinician / patient)

• Pathologists

• Demographics (race, gender, age differences in presentation)

• AI training bias (trained on limited populations e.g., young white men)

• Risk of missing older women / minority ethnic groups

• Diet influencing biopsy presentation (e.g., low-inflammation diets)

• Press / media

• Patients

• Coeliac UK

• Funding bodies

• NHS

• Venture capitalists

• Funders (ROI focus)

• NHS commissioners

• Healthcare professionals

• GPs

• Consultants

• Patients' care team

• Government health departments

• Politicians (funding decisions)

• Health trusts / boards

• NHS regulators

• Ethical council

• Data providers

• Clinicians

• General public

• People with undiagnosed symptoms

• Stakeholders (e.g., GP / consultant funders)

• NHS system itself

• Patients + families

• Confidence in AI dependent on follow-up outcomes

• AI outperforming pathologists (where disagreement exists)

• AI identifying disease / normal gut more accurately than pathologists

**How might we do that?**

• Demonstrate results

• Have a safety net

• Evidence from trials

• Prove accuracy

• Proof of accuracy

• At least as good as pathologists (or better)

• Blind testing vs pathologists

• Double blind comparison (AI vs pathologists)

• Train AI on cases where pathologists disagree

• Identify what AI misses

• Standardised verification process

• Controls against human comparison

• Ongoing validation (over time)

• Large, diverse dataset for training

• No discrimination (all cases detected)

• Demonstrate reliability at scale

• Case studies

• Research outputs published

• Peer review of methodology

• Data on accuracy

• Evidence of success rates

• Figures shared with patients

• Inconclusive results double-checked by pathologist

• Second opinion option if symptoms persist

• Further confirmatory test after AI diagnosis (e.g., biopsy)

• Set standard dye mix for AI biopsy test

• Quantify benefits

• Cost saving evidence

• Cost–benefit analysis

• ROI for funders

• Faster and cheaper than current pathway

• Increased speed of diagnosis

• Extend AI to other conditions (increase ROI)

• Show how funding will work

• Educate GPs

• Target GPs and gastroenterologists

• Information sessions

• Transparency with care providers

• Explain safeguards

• Explain humans are not 100% accurate

• Reassurance about correct data input

• Data security (not hacked)

• Ongoing monitoring and updates

• Patient-facing information (easy-read if requested)

• Use patient stories

• Use Coeliac UK

• Ride on other successful AI deployments

• Work with other organisations (e.g., Osteoporosis Society)

• Engage Scottish Health Minister / Westminster

• Political support for funding

• Demonstrate societal benefit (e.g., fewer sick days)

• Test on volunteers (established coeliac patients)

• Re-testing after period of time

• No need to be on gluten diet prior to testing (clarification required)

• Make features / model explanation clear

**Where are the potholes? (risks to our strategy and suggested improvements)**

• Will method work in other countries (different diets e.g., lower gluten / Mediterranean / sub-Saharan / Indian)

• Diet influencing biopsy presentation

• Include different races in training data (e.g., BAME groups)

• Concern AI “not tested on us” (vaccination comparison)

• Data continues to recognise different demographic groups accurately

• Training implications for new pathologists

• Skill fade in experienced pathologists

• AI replacing learning opportunities

• Still require human pathologist oversight

• Who provides second opinion?

• Lack of pathologists to review results

• AI = vague / confusing term (deepfake association)

• Public misunderstanding of AI

• Negative assumptions / prejudices about AI

• Patients refusing AI-based diagnosis

• People's trust

• Do not overstate benefits to end users

• Explain clearly how system works

• Part of diagnostic toolkit (not sole decision-maker)

• AI hallucinations

• “Computer says no” concern

• Matching AI + pathologist opinion could still be wrong

• Accuracy / reliability concerns

• False negatives prolonging diagnosis

• Likelihood score vs binary output

• Indeterminate outcomes require human review

• Regular spot checks (algorithm drift)

• Validation of test criteria

• If no objective gold standard, how measure accuracy?

• Information loss from stain standardisation

• Robustness of AI model

• Re-biopsy to confirm improvement (time + cost)

• Delay waiting for biopsy

• Bottleneck at biopsy procurement

• Ability to increase biopsy capacity

• Nothing changes if biopsy numbers unchanged

• Second biopsy adds time and expense

• Cybersecurity risks

• Cyberattacks

• Malware vulnerability

• Single point of failure

• Leaked privacy / metadata

• Data security

• Data storage duration

• Verification and maintenance

• Data / power requirements

• Volume of data required

• Large-scale validation

• Centralised vs distributed delivery model

• Technology costs (processing, storage)

• Equipment change costs

• Health board willingness to invest

• Funding for rollout

• NHS funding constraints

• Healthcare devolved across UK nations

• Compare international testing models (e.g., Italy)

• Ethical considerations

• Waiting until patients are very ill before testing

• NHS funds

• Public opinion

• Discussion groups too large / noisy

• AI-enabled non-invasive alternatives (scan gut instead of biopsy)

**Additional individual comments (facilitator-recorded)**

• “In favour, it's already being used in radiology, it's faster and my experience has been good. But it's not solely completed by a machine, a human is involved in the process.”

• “I feel there is misuse of labelling things with AI as it is mainly associated with bad things. I think machine learning automation is much better.”

• “I don't think it should fully replace a human and what about cyber attacks tampering with the data, what about security?”

• “100% agree I would have someone revisit my biopsies.”

• “I had two GP misdiagnoses, so why not give AI a go!”

• “If there is a difference in opinion between AI and pathologist I would want a third opinion.”

• “Sharing of full data between primary care and secondary care is not in place and that is a problem.”

• “Biopsy should not be in isolation; if there is disagreement then a human should step in and make a decision based on all the available information.”

• “Machine learning [is] more acceptable to the younger generation” (older participant); “my generation don't have a problem with it” (younger participant).

• “You will need to lobby the politicians to accept this, they will need to know the savings to the NHS.”

#### Supplementary Table 7

**Cambridge PPIE workshop (Hughes Hall, June 2026) - raw discussion notes as recorded by facilitators (anonymised)**

***Presentation Q&A (before focus groups)***

**Notes by first note-taker**

**Questions asked at the end of the presentation**

• 1. How do you know the specificity of the AI, if the biopsies it's being trained on don't have a diagnosis that pathologists agree on?

Retrospectively compare to historic diagnosed CD cases

• 2. How do you eliminate ethnicity, gender and age biases?

Tricky to do but can try to eliminate some bias by ensuring the data set is very large and that it contains biopsies from people across wide demographics. Using samples from real world hospitals so although there may be some bias, it's being trained to align with real world practise. Epidemiology of CD shows there are fewer people from diverse backgrounds with CD / with a formal diagnosis and therefore there are less samples available and ethnicity is not recorded well. Saw an issue with paediatric samples on the system initially so added more samples to the study. Accuracy on paediatric samples is important for future screening.

• 3. What happens if biopsy is disrupted?

Will be okay, as often they are but can still be viewed in current practise so it would be the same, only not possible if not at all viable.

• 4. Can it look at all biopsies or just those suspected to be CD?

Can look at all as trained to identify and push non-CD cases appropriately

• 5. How do you ensure other diagnoses don't get overlooked?

Only certain biopsies will be put into the algorithm, e.g., those not likely to be cancer based on clinical suspicion. Those which are put into the system can be flagged as definite CD, uncertain and requiring pathologist view, other conditions would fall into this and be viewed.

• 6. If 75% of all biopsies are normal, can it be used for reducing the work load?

Yes

• 7. If flagged as not CD, does it answer the question it came with?

[Noted by facilitators as a question for further consideration]

• 8. Are there national guidelines for pathology for CD?

Not hard and fast

• 9. Have you done any time saving modelling?

Yes, but only 'back of envelope' at this stage. Depending on uptake it could save up to 10 minutes per biopsy of pathologist time but won't save time from referral to actually getting the biopsy done.

• 10. Due to accuracy and speed will it reduce the number of people who die without ever getting a diagnosis?

Potentially, but people still need to get through the door and on the diagnosis journey

• 11. Can my biopsy samples now be looked at with the system?

Yes – something we'd like to offer in theory

**Notes by second note-taker**

• Presentation Q&A: One participant asked about the concerns of pathologists in the industry, which led to discussion about potential deskilling of junior pathologists which the group was understanding of.

• One other participant asked about how potential diagnoses may be sidelined if a multimorbid biopsy reveals additional diagnoses in addition to coeliac, and fail-safes for this instance.

• There were concerns over other diagnoses that could resemble features of coeliac disease (e.g., GvHD, tropical sprue).

**Focus Group Notes**

**Notes by third note-taker**

• To the question of whether they're using AI in everyday life: can't avoid AI these days (e.g., when googling things, the first thing coming up is an AI overview). Several people said they're trying to minimise it (Google example, scroll past AI), but hard to do.

• One person said (and several agreed) that they were worried about the impact of AI on climate change, e.g., even if a medical AI software improves care in the UK, would the increased carbon emissions have a bigger negative effect somewhere else in the world.

• Another person replied saying that the key is to know about the carbon emissions, because it's possible that there may be a net benefit (e.g., AI using fewer carbon emissions than pathologists).

• Generally low confidence in public AI models (e.g., chatbots), because it is not known where they got the data from, how it was trained etc., leading to lower confidence.

• About LLMs, one person said “garbage in, garbage out”; however, with our AI they thought it was the right tool for the right job. For Lyzeum, like in healthcare more generally, the training dataset is very important, but they found it reassuring that a lot of focus had been put into it.

• Regulation of medical devices gives you confidence.

• Medical AI is self-contained (answering one specific question), leading to much larger trust than a general purpose LLM.

• One person was worried about data security (worried that information would get hacked/leaked).

• One person was worried about bias of the AI.

• Two people agreed that healthcare is the ideal application for AI, as it is solving one very specific purpose.

• Several people felt much more confident with algorithms that were owned and controlled by UK companies/groups. They felt very different about UK AI compared to international companies having access to our data (Palantir was given as an example of something they disapproved of).

• One person was worried that AI would increasingly turn the UK into a 2-tier healthcare system, e.g., what happens to the NHS – will the software be used in all NHS hospitals or only some, and might the NHS not pay for it at all while private healthcare providers do.

• Several people mentioned that they thought it was “mad” that you couldn't share data between NHS trusts. They thought the fragmentation should be improved, so that it's easier for training and evaluation of AI. One person told a story of having an endoscopy for something non-CD related, where a biopsy for coeliac disease was also taken because the team didn't know the patient had already been diagnosed with CD in a different trust. Someone else told a story of having to be the messenger between two trusts as they couldn't communicate directly. They also thought it would be very hard to overcome barriers between hospitals.

• Someone questioned the way hospitals were being evaluated, giving an example of Addenbrooke's hospital installing a new technological system which performed very well but was a bit more expensive, so it lost a lot of “points”.

• On what would happen if the AI disagreed with the pathologists: one person said they would trust whichever provided a diagnosis that fits with the patient's narrative based on the symptoms they experience, e.g., one person said they would just try a gluten-free diet and see whether that helps.

• People talked about differences in symptoms: one person was anaemic but never had any other symptoms; another had negative blood tests for five years, but their biopsy was then CD positive.

**Notes by second note-taker**

How do you use AI in daily life? If you don’t use it, what stops you from using it?

• One participant works as a development manager for a SaaS software company. Mentioned new trend of vibe coding.

• Concerns about inaccuracy of AI in Google searches, and preference to see data and patterns. Concerns of implicit bias and mentioned the book ‘Weapons of Math Destruction’.

How do you think AI might be useful in healthcare? Do you have any concerns around the use of AI in healthcare?

• Participant who works in healthcare uses AI for scribing, which helps clinicians have more time with their patients and improves patient experience.

• Another participant mentioned how GP appointments have the same tools, especially for generating referral letters.

• Concerns about people using AI to self-diagnose.

• Concerns about AI’s confidence and agreeability.

• Concerns about how well regulated this really will be, and how the public will know of these details.

• Concern about losing the nuance of the patient record to support accuracy.

• Mention of centralised vs. individual trust hosting the server for Lyzeum AI.

How would you feel about a regulated programme which lets you put your symptoms in and give you a diagnosis?

• Needs to be made very clear that it’s AI.

How do you think people might react if a computer system gave a different diagnosis to one given by a trained histopathologist in the past? (i.e. retrospective)

• Re-conduct test as there is an element of doubt.

• Perhaps age may be an influence; an older patient may trust the pathologist more, whilst a younger patient may trust the AI more.

• Might use symptoms to help determine which is more accurate; but this is made harder given that coeliac disease does not always present symptoms (but they are still a coeliac disease positive patient).

**Notes by fourth note-taker**

How do you use AI in daily life? If you don't use it, what stops you from using it?

• AI used by all members of the group to varying extents and purposes, typically chatbots (ChatGPT, Gemini) in the place of Google for sourcing information (e.g., identifying plants when gardening).

• Two software engineers present in the group used AI for work/coding tasks but “take it with a pinch of salt” and use professional discernment when looking at outputs (tweaking prompts, highlighting inaccuracies). They expressed that AI was too broad or potentially misleading a term when used in a clinical context, and agreed that “machine learning” was a more appropriate description of the technology.

• All members made the point of highlighting how heavily integrated AI is into popular search engines by default, and reiterated that its outputs and summaries aren't always accurate and shouldn't be accepted blindly.

How do you think AI might be useful in healthcare? Do you have any concerns around the use of AI in healthcare?

• General consensus that AI is a useful tool that must be used responsibly, especially in healthcare settings. All agreed that an AI is only as good as the quality of data used to train it (“garbage in, garbage out”).

• All members felt that AI developed in a similar way to Lyzeum's would be useful and trustworthy.

• Concerns expressed about AI being used in healthcare to cut costs (seen as both a positive and negative depending on individual interpretation), and with it the increased risk or opportunity of greed and subsequent corner-cutting/low-quality results at the expense of patients.

• Additional concerns raised regarding the extent of human oversight, transparency during patient appointments (i.e. allowing providers to show the AI's justifications for its decisions, such as crypt-villi measurements and IEL count), and the critical need for a human at the end of the process to provide emotional support and empathy.

How do you think people might react if a computer system gave a different diagnosis to one given by a trained histopathologist in the past?

• All members of the group had a generally positive outlook on diagnostic AI/computer systems (especially if they functioned similarly to Lyzeum's), and would trust it over a trained pathologist if a conflicting diagnosis was made.

• A few members even expressed interest in having their own past biopsies and clinical information re-evaluated using Lyzeum's AI, repeatedly citing the low level/percentage of agreement between pathologists when forming a diagnosis as a key reason for feeling this way.

**Notes by first note-taker**

1. How do you use AI in daily life? If you don't use it, what stops you from using it?

• A number of people shared that either they exclusively or most commonly used Google Gemini, as it has a feature that automatically provides answers and/or information with references to everything searched via Google's search engine, their preferred search engine of choice.

• A number of people shared that they used ChatGPT for various tasks in both a work setting and a non-working, personal setting: drawing out the key points of a long contract or legal document, summarising long documents, improving grammar/spelling/writing style, and pulling out and explaining technical language.

• Two individuals reported using AI to develop resources for lessons in school or the workplace that they themselves could then adapt and build upon – only using AI to get them started, then improving it, checking references, improving tone, and removing Americanisms.

• A few people commented that they only used AI for trivial things and wouldn't rely on it for something important, e.g., finding answers to silly questions quickly (“how many bouncy balls would it take to fill planet earth?”).

• A now-retired chief in the oil industry shared that their role required them to use AI to support data processing, introduced around 2017.

• A now-retired breast cancer radiographer shared that AI was introduced to support healthcare professionals in identifying abnormalities and homing in on areas for further investigation.

• A parent shared that they'd used AI to interpret blood test results in the long wait between results being uploaded to the NHS app and being able to see a clinician to discuss them.

• Two individuals in the group were openly against using AI in their personal life. One cited environmental reasons (the systems require a large amount of power, which is not sustainable long-term); the other cited wariness about data handling and safeguarding measures.

2. How do you think AI might be useful in healthcare? Do you have any concerns about the use of AI in healthcare?

• A few members agreed it could be very useful in translating health information into layman's terms to improve patient understanding.

• A few members felt it could be used to support symptom checkers, though this could have positive and negative impacts. Positive impacts discussed: better awareness of conditions, earlier contact with healthcare professionals, earlier diagnosis and better health outcomes. Negative impacts discussed: ‘more hypochondriacs’, increased worry, and increased strain on healthcare services from people seeking help where they may not have otherwise.

• One person suggested it could be used to look at patient data and find links between conditions, flag risk factors, identify patterns or problems, and support research into condition development or comorbidities. Another suggested it could be used to look at health information retrospectively to look at trends in health and suggest possible solutions, e.g., obesity. It could do jobs that humans do not have the time or capacity to do in a much shorter timeframe, and could flag to a GP that a person should have a certain test based on key symptoms.

• The group agreed a key benefit could be reducing health inequalities and human bias, and that it could relieve strain and help solve staffing shortages and systemic issues across the NHS, though training staff on new systems is time-consuming and has to be done properly to be effective.

• Concerns included worries about the safety of data (particularly sensitive health data), lack of informed consent, and external contracts with processors and server companies. It was noted that audits, regulations and regular checks would mitigate some of these issues; one person noted that tech regulation is often behind technology as it evolves, a particular concern given AI's rapid pace of development. It was also noted that new healthcare professionals would need to be properly trained in the context of AI being part of their role.

• Two individuals explicitly stated that they felt human consultants should always get the final say regardless of the system used, and that AI should only be an aid – citing the value of bedside manner and human connection, and greater confidence in humans over systems.

3. How do you think people might react if a computer system gave a different diagnosis to one given by a trained histopathologist in the past?

• A number of people stated that they didn't care how the diagnosis was given, they just wanted it to be accurate – so if a new diagnosis process (AI or otherwise), proven by research to be more accurate than the previous method, gave a different diagnosis, they would accept it.

• Some people reported it would depend on symptoms: if they had felt better after going gluten-free following a diagnosis, they would be less inclined to believe a new diagnosis stating they maybe did not have coeliac disease.

• A number of people agreed they would require a third opinion, maybe even a fourth, but that this would be the case even if another human gave a different opinion.

• One person reported that they would always want to consult a human.

**Notes by fifth note-taker**

1. How do you use AI in daily life? If you don't, what stops you from using it?

• One attendee, a teacher, shared that they used AI predominantly to communicate with parents (checking the best wording) and to prepare and take notes.

• Another attendee, also a teacher, shared negative views on AI, feeling it should be avoided in a school setting. They thought it was useless and provided incorrect information on many topics, and that seeing students use it prevents them from correctly learning information and thinking for themselves. There was agreement that AI felt like the lazy option in many scenarios.

• Other attendees agreed that the information AI provides can be incorrect and out of date.

• One attendee said they thought the form of AI mainly used (free online sources like ChatGPT) was very different from the machine learning described in the presentation, and felt it was difficult to distinguish between the two in the questionnaire; there was some agreement on this within the group.

• Social media was said to be a big aspect of encountering AI in daily life, and it was felt to be difficult to distinguish what was true information.

• One attendee shared that they use AI for everyday research purposes as part of their job role, favouring paid accounts over free online AI chatbots because they seemed more secure and trustworthy. While they could see why people are pushing for AI and its possible benefits, they remained sceptical and felt caution needs to be taken with its use – though they thought it had the best applications for medicine.

• Another attendee, who works in publishing, sees AI being used often, is sceptical about its use, and feels it disregards authors' voices, seeing it as a possible worry for job security in that sector.

• Despite these concerns, the group noted that humans can also make errors.

• The group agreed AI was better used as an “advanced Google”, most useful when someone with the relevant expertise picks out the useful and correct information; the greater danger was people blindly believing all information.

2. How do you think AI might be useful in healthcare? Do you have any concerns?

• One attendee shared that they had initial concerns about AI in healthcare before attending the workshop, but felt the presentation had improved their feelings toward it; a few others in the group agreed.

• A few attendees agreed they felt less worried knowing that pathologists were still involved in the diagnosis pathway.

• One attendee mentioned the importance of training the AI tool on a wide portfolio of data, including across genders and ethnicities to diminish bias, and highlighted the need for consent to use patients' data.

• It was widely agreed that AI would be most effective for easier tasks, saving pathologists time and reducing excess pressure. A few attendees also supported using AI to set up and schedule appointments; however, one attendee felt AI was less useful for writing up patient 1-1 appointments, as although it would save time, it wouldn't pick up non-verbal indicators.

3. How do you think people might react if a computer system gave a different diagnosis to one given by a trained histopathologist in the past?

• Most attendees agreed that some people are desperate to be diagnosed and have an answer to their symptoms, and would therefore be inclined to follow whichever answer gave them a coeliac disease diagnosis.

• Attendees agreed it would need to be handled sensitively.

• A couple of attendees said that if they had previously been diagnosed with coeliac disease by a trained histopathologist and a computer system then told them they did not have coeliac disease, it would not change their lifestyle – they felt they knew how gluten affected them and would remain gluten-free regardless.

• One attendee felt that if a previous positive diagnosis had been invalidated by a computer system, it may make it more difficult to follow a gluten-free diet, explaining it might mean their diet requirements weren't taken as seriously and their access to support and follow-up scans (particularly DEXA bone scans) would be limited.

• A few members agreed it would be a different scenario for a cancer diagnosis, since delaying a coeliac disease diagnosis is not as detrimental as delaying some cancer diagnoses.

• One attendee said they would be unhappy and feel invalidated if they received a negative result from a computer system after a previous coeliac disease diagnosis, and would not seek a new biopsy for re-testing as they would not be able to eat gluten.

• There was worry for people who had a previously negative biopsy but where the computer system then found a positive result – fear centred on possible complications of having eaten gluten in the interim, dependent on the timeline, symptom severity, and whether the patient was a child.

• One attendee felt that any mention of coeliac disease already offered a possible solution – regardless of the final diagnosis, the mention opened up a gluten-free diet as an avenue to see if it helped improve symptoms.

**Notes by sixth note-taker**

How do you use AI in daily life and if you don't, what stops you?

• “I use open AI coding for work that is being used in healthcare, I believe it is saving me time, maybe!”

• “I use ChatGPT on my phone to ask it to interpret my blood test results and give me an answer in lay terms.”

• “I use Google AI, and its answers are generally good. However, when I already know the answer, I can see that it is sometimes incorrect. It's a catch 22 – if I don't already know the real answer, how will I know what it's telling me is right or wrong! AI ancestry is still quite poor, although it is improving. I'm not totally confident.”

• “I use search assist, but I am wary AI is still learning.”

• “I don't think AI will give any credibility to ancestry. I've not used ChatGPT but I am impressed with some things I've seen, like today. I use co-pilot a reasonable amount. I have asked about plants and, as this is my area of expertise, I would estimate AI is correct about 80–90% of the time.”

• “I am a software engineer and I find AI can speed things up, increase productivity – it's easier than searching a help function – but there are risks that need to be interpreted.”

How do you think AI might be useful in healthcare? Do you have any concerns around the use of AI in healthcare?

• “The example today! A useful tool, it cuts out the donkey work which frees up time for other work where AI can't be used. It's ok to use AI if it has been proven to be valid/trustworthy.”

• “In today's example it's ok to use AI so long as borderline cases are still referred to a human.”

• “I have a concern regarding cybersecurity, links in the chain increase risk.”

• No one in the group had any previous experience of the use of AI in healthcare when asked (although later in the discussion, someone referred to using a GP bot).

• “In the case of a busy GP they may not suspect a cause, but an AI machine-learned system may detect and propose a possible solution.”

• “An AI tool should be used by human beings, not instead of human beings, and targeted to improve patient outcomes, not for reducing costs – need to have the correct motivations!”

• “Agree it shouldn't be about saving money but focused on improving productivity and patient outcomes.”

• “I think it would be important to understand the mathematics and probabilities of the output.”

• “Thinking about the example this morning, would there be some ethnicities that would be missed by this AI system? If it hasn't been trained with biopsies from those ethnicities, would you need to target groups as part of the learning process?”

• “I have used a GP bot to make an appointment and it's not a good experience, as it removes all the emotional understanding.”

How do you think people might react if a computer system gave a different diagnosis to one given by a trained histopathologist in the past?

• “I think I would now be doubtful of the histopathologist result and would trust AI after what I have heard today.”

• “Same, I agree after seeing the presentation today.”

• “I would want to sit down with my GP and have a conversation, but I would be worried the NHS would not provide that opportunity!”

• “I'd have more confidence in the AI result.”

• “I would want a full explanation of the AI results, e.g., villi, crypts, IELs – this is the spectrum and your results sit here in the spectrum. That way I would have greater context.”

• “AI, once trained, will be consistent in its output, whereas a healthcare professional is a human being – they will have off days, good days, bad days, and may work under par, but the AI won't be affected by that.”

• “I don't mind the use of AI, providing a human makes the final assessment/interpretation.”

#### Supplementary Table 8

**Cambridge PPIE workshop - pre-session questionnaire (Questionnaire 1), full quantitative results (n=34)**

Responses collected immediately before the structured educational session at the Cambridge PPIE workshop (June 2026), prior to matched post-session responses reported below.

**Demographics**

*Age: mean 58.7 years, median 62.5, range 23–84 (n=32 valid numeric responses of 34 answered; 2 participants did not disclose their age).*

**Gender**

*What is your gender?*

| **Response option** | **n** | **%** |
| --- | --- | --- |
| Male | 13 | 38.2% |
| Female | 21 | 61.8% |
| Prefer not to say | 0 | 0.0% |
| Other (please specify) | 0 | 0.0% |

**Education**

*What is your highest level of education?*

| **Response option** | **n** | **%** |
| --- | --- | --- |
| Left school at 16 | 4 | 11.8% |
| Left school at 18 | 3 | 8.8% |
| Undergraduate degree | 10 | 29.4% |
| Postgraduate (Masters) | 8 | 23.5% |
| Postgraduate (PGCE) | 1 | 2.9% |
| Postgraduate (Masters + PGCE) | 1 | 2.9% |
| Postgraduate (PhD/Doctorate) | 5 | 14.7% |
| Postgraduate (Unspecified) | 2 | 5.9% |

**Family history of CD**

*Have you or a family member been diagnosed with coeliac disease?*

| **Response option** | **n** | **%** |
| --- | --- | --- |
| Yes | 33 | 97.1% |
| No | 1 | 2.9% |

**Personal biopsy history**

*Have you ever had an upper GI biopsy taken?*

| **Response option** | **n** | **%** |
| --- | --- | --- |
| Yes | 31 | 91.2% |
| No | 3 | 8.8% |

**Worked in healthcare**

*Have you ever worked in healthcare?*

| **Response option** | **n** | **%** |
| --- | --- | --- |
| Yes | 6 | 17.6% |
| No | 28 | 82.4% |

**Worked in technology or AI**

*Have you ever worked in technology or AI?*

| **Response option** | **n** | **%** |
| --- | --- | --- |
| Yes | 6 | 17.6% |
| No | 28 | 82.4% |

**Attitudes to AI (Likert agreement)**

**Positive about AI in general**

*I feel positively about artificial intelligence in general.*

| **Response option** | **n** | **%** |
| --- | --- | --- |
| Strongly disagree | 2 | 5.9% |
| Disagree | 6 | 17.6% |
| Neither agree nor disagree | 12 | 35.3% |
| Agree | 14 | 41.2% |
| Strongly agree | 0 | 0.0% |

**Trust in AI in healthcare**

*I trust the use of artificial intelligence in healthcare.*

| **Response option** | **n** | **%** |
| --- | --- | --- |
| Strongly disagree | 1 | 2.9% |
| Disagree | 1 | 2.9% |
| Neither agree nor disagree | 19 | 55.9% |
| Agree | 12 | 35.3% |
| Strongly agree | 1 | 2.9% |

**AI could improve CD diagnosis**

*Artificial intelligence could improve the diagnosis of coeliac disease.*

| **Response option** | **n** | **%** |
| --- | --- | --- |
| Strongly disagree | 0 | 0.0% |
| Disagree | 1 | 2.9% |
| Neither agree nor disagree | 15 | 44.1% |
| Agree | 14 | 41.2% |
| Strongly agree | 4 | 11.8% |

**Comfort with AI-assisted diagnosis**

*I would feel comfortable if artificial intelligence assisted doctors in diagnosing coeliac disease.*

| **Response option** | **n** | **%** |
| --- | --- | --- |
| Strongly disagree | 0 | 0.0% |
| Disagree | 0 | 0.0% |
| Neither agree nor disagree | 5 | 14.7% |
| Agree | 23 | 67.6% |
| Strongly agree | 6 | 17.6% |

**Importance of AI explainability**

*It is important that medical artificial intelligence systems can explain how they make decisions.*

| **Response option** | **n** | **%** |
| --- | --- | --- |
| Strongly disagree | 0 | 0.0% |
| Disagree | 0 | 0.0% |
| Neither agree nor disagree | 2 | 5.9% |
| Agree | 13 | 38.2% |
| Strongly agree | 19 | 55.9% |

**Retrospective views (preceding 2 years)**

**Retrospective view: AI in general (2yr)**

*Over the past 2 years, my view of artificial intelligence in general has become…*

| **Response option** | **n** | **%** |
| --- | --- | --- |
| Much more negative | 4 | 11.8% |
| Slightly more negative | 6 | 17.6% |
| Stayed the same | 7 | 20.6% |
| Slightly more positive | 13 | 38.2% |
| Much more positive | 4 | 11.8% |

**Retrospective view: AI in healthcare (2yr)**

*Over the past 2 years, my view of artificial intelligence in the delivery of healthcare has become…*

| **Response option** | **n** | **%** |
| --- | --- | --- |
| Much more negative | 1 | 2.9% |
| Slightly more negative | 0 | 0.0% |
| Stayed the same | 11 | 32.4% |
| Slightly more positive | 18 | 52.9% |
| Much more positive | 4 | 11.8% |

**Cambridge PPIE workshop - post-session questionnaire (Questionnaire 2), full quantitative results (n=34)**

Responses collected immediately after the structured educational session at the Cambridge PPIE workshop (June 2026), matched to the pre-session responses reported above.

**Attitudes to AI (Likert agreement)**

**Positive about AI in general**

*I feel positively about artificial intelligence in general.*

| **Response option** | **n** | **%** |
| --- | --- | --- |
| Strongly disagree | 3 | 8.8% |
| Disagree | 1 | 2.9% |
| Neither agree nor disagree | 13 | 38.2% |
| Agree | 15 | 44.1% |
| Strongly agree | 2 | 5.9% |

**Trust in AI in healthcare**

*I trust the use of artificial intelligence in healthcare.*

| **Response option** | **n** | **%** |
| --- | --- | --- |
| Strongly disagree | 0 | 0.0% |
| Disagree | 1 | 2.9% |
| Neither agree nor disagree | 10 | 29.4% |
| Agree | 19 | 55.9% |
| Strongly agree | 4 | 11.8% |

**AI could improve CD diagnosis**

*Artificial intelligence could improve the diagnosis of coeliac disease.*

| **Response option** | **n** | **%** |
| --- | --- | --- |
| Strongly disagree | 0 | 0.0% |
| Disagree | 0 | 0.0% |
| Neither agree nor disagree | 1 | 2.9% |
| Agree | 13 | 38.2% |
| Strongly agree | 20 | 58.8% |

**Comfort with AI-assisted diagnosis**

*I would feel comfortable if artificial intelligence assisted doctors in diagnosing coeliac disease.*

| **Response option** | **n** | **%** |
| --- | --- | --- |
| Strongly disagree | 0 | 0.0% |
| Disagree | 0 | 0.0% |
| Neither agree nor disagree | 1 | 2.9% |
| Agree | 16 | 47.1% |
| Strongly agree | 17 | 50.0% |

**Importance of AI explainability**

*It is important that medical artificial intelligence systems can explain how they make decisions.*

| **Response option** | **n** | **%** |
| --- | --- | --- |
| Strongly disagree | 0 | 0.0% |
| Disagree | 0 | 0.0% |
| Neither agree nor disagree | 0 | 0.0% |
| Agree | 9 | 26.5% |
| Strongly agree | 25 | 73.5% |

**Views at end of workshop**

**View by end of workshop: AI in general**

*At the end of this workshop, my view of artificial intelligence in general has become…*

| **Response option** | **n** | **%** |
| --- | --- | --- |
| Much more negative | 0 | 0.0% |
| Slightly more negative | 0 | 0.0% |
| Stayed the same | 10 | 29.4% |
| Slightly more positive | 19 | 55.9% |
| Much more positive | 5 | 14.7% |

**View by end of workshop: AI in healthcare**

*At the end of this workshop, my view of artificial intelligence in the delivery of healthcare has become…*

| **Response option** | **n** | **%** |
| --- | --- | --- |
| Much more negative | 0 | 0.0% |
| Slightly more negative | 0 | 0.0% |
| Stayed the same | 2 | 5.9% |
| Slightly more positive | 17 | 50.0% |
| Much more positive | 15 | 44.1% |

**Workshop evaluation**

**Workshop was well organised**

*The workshop was well organised.*

| **Response option** | **n** | **%** |
| --- | --- | --- |
| Strongly disagree | 0 | 0.0% |
| Disagree | 0 | 0.0% |
| Neither agree nor disagree | 0 | 0.0% |
| Agree | 7 | 20.6% |
| Strongly agree | 27 | 79.4% |

**Enjoyed attending the workshop**

*I enjoyed attending the workshop.*

| **Response option** | **n** | **%** |
| --- | --- | --- |
| Strongly disagree | 0 | 0.0% |
| Disagree | 0 | 0.0% |
| Neither agree nor disagree | 0 | 0.0% |
| Agree | 9 | 26.5% |
| Strongly agree | 25 | 73.5% |

**Felt voice was heard and valued**

*I felt my voice was heard and my contribution was valued.*

| **Response option** | **n** | **%** |
| --- | --- | --- |
| Strongly disagree | 0 | 0.0% |
| Disagree | 0 | 0.0% |
| Neither agree nor disagree | 0 | 0.0% |
| Agree | 11 | 32.4% |
| Strongly agree | 23 | 67.6% |

#### Supplementary Table 9

**Cambridge workshop (6 June 2026) - Questionnaire 2 full free text responses (34 completed questionnaires)**

**Q3: What, if anything, specifically changed your view? (or, if unchanged, what might have changed it)**

• EXPLANATIONS ABOUT HOW MACHINE LEARNING WORKED AND WAS TRAINED, VERY HELPFUL.

• The system described is based on clear biological and quantifiable factors. Decisions are thus more clearly defined than human interpretation.

• The machine learning is very specific and the use of AI to assist diagnosis can be evidenced leading to more confidence.

• Better understanding.

• Speeds up processing. Seeing how it works and the benefits has made me more positive.

• I feel more positive about AI. Explanation about process + checks involved.

• I think my view has become more positive.

• Pathology 75% success/agreement - AI 97%.

• I know about AI use in diagnostics but for coeliac disease it is very different. I am aware that AI won't go away but I am reassured checks are in place and control of outputs.

• Using the language “Machine learning”. Knowing that the data has been checked against patient outcomes gives me more confidence.

• MY ORIGINAL ANSWER WAS BASED ON MY LACK OF KNOWLEDGE REGARDING AI. AFTER THE PRESENTATION I FEEL MORE CONFIDENT IN THE USE OF AI.

• It's good to hear about the controls + regulation for Medical ML. It's very different from the LLM wild west and I think that isn't obvious to people.

• Rigorous regulation.

• MY VIEW OF AI IN GENERAL (I.E. PUBLICLY AVAILABLE AI) REMAINS NEGATIVE BUT THE APPROACH TO THIS PROJECT HAS OUTLINED HOW AI CAN BEST BE USED AND CONTROLLED IN THE HEALTHCARE ENVIRONMENT.

• Machine learning on images is a narrow subset of AI and I have more confidence in this than generative AI. Accuracy >95% vs pathologist agreement ~80% seems clear improvement.

• I am impressed by the AI advances and the pathologists' care.

• I FOUND THE PRESENTATION VERY CONVINCING… I HAD BEEN MISDIAGNOSED (OR RATHER NOT DIAGNOSED) BECAUSE OF OVERRELIANCE ON BLOOD TEST (TO WHICH I AM NEGATIVE) AND WHEN I HAD ENDOSCOPY, EMPHASIS WAS ON POSSIBILITY OF CANCER.

• A member of the research team's description of how machine learning learns to spot coeliac disease was reassuring, as previously I found it difficult to understand or feel confident in the way it arrived at the output results.

• AI hugely positive in healthcare because intelligent, responsible people are controlling it. AI not positive when in the hands of those who aren't intelligent or responsible.

• Understanding how AI could be useful.

• Superb and convincing presentation on this healthcare project. General AI - climate worry, power of tech billionaires also thoroughly alarming for democratic systems - understandably this session didn't address the wider issues, and they do not, after all, really impact on the project. Welcomed the commitment to record energy usage involved.

• The presentation explained the way the tool could improve diagnosis and reduce the pressure on the NHS.

• I feel more positive having heard about the rigorous tests and the regulatory framework.

• The regulation already around it. Ensuring the data doesn't have biases.

• I think learning how decisions are made makes me more confident -> the AI decisions.

• My view might have changed if there was very clear guidance that it would save significant time in the overall diagnostic process but I'm not sure the time saving offsets potential downsides.

• Understanding the checks and balances that have contributed to the process.

• Understanding the safeguards and approval process.

• Hearing about the process, data has been very useful.

• Lots of information given which helped.

• If too many differing results from AI to physical exam then probably yes.

**Q4: Are there any further comments, concerns, or suggestions you would like to share about AI in healthcare or coeliac disease diagnosis?**

• AI will remove lower level and simple processes so how will training take place.

• PRESUMABLY THE SAME TYPE OF MACHINE LEARNING CAN BE APPLIED TO OTHER HISTOPATHOLOGY DIAGNOSES?

• I would be happy with this system.

• Concern about rigidity and inability of AI to adapt to change.

• No.

• Would like to be sure that once AI process in place there would still be checks as time went on.

• I think there should be more workshops on the subject.

• I hope it enables Pathologists/Clinicians to focus on more complex cases / aspects of work load.

• Gender biases.

• I like the approach of normal vs coeliac vs everything else.

• No.

• Good luck with it!

• UNDERLYING CONCERN OVER THE INTEGRITY OF THE MODEL I.E. MANAGING ACCIDENTAL / DELIBERATE DATA ERRORS.

• Concern about cybersecurity and dependence of pathology (systems hack).

• But I do value especially and trust professional human final opinion.

• PLEASE REVIEW OVERRELIANCE OF BLOOD TEST DIAGNOSIS OF COELIAC DISEASE - I WAS ONLY DIAGNOSED PRIVATELY AFTER YEARS OF SYMPTOMS.

• Slightly concerned in the lack of agreement between pathologists currently and what this will mean if they are then responsible for diagnosing the more complex cases that AI is unable to diagnose confidently.

• It needs to be made very clear there is a difference between the use of generative LLMs & machine learning.

• Interesting points in our roundtable discussion. A member of the research team took notes (as well as giving us wonderfully informative responses).

• As we discussed in our group, I would be keen to learn about the tool (or other tools) which can diagnose coeliac when you have been following a gluten free diet already.

• I think clear explanation of process and benefits is key.

• I feel this is a great application of AI in general. If it can save time and money + lead to more correct diagnosis - that is a win!

• AI in healthcare - concern about level of scrutiny of private AI companies then having access to sensitive healthcare data. Strong concerns about environmental impact of AI regardless of the sector it is used in.

• Future job security.

• Rules, Regs & Data etc re Coeliacs.

• None.

#### Supplementary Table 10

**Healthcare professional survey (Lyzeum Ltd) - full quantitative results (n=37 respondents; item-level n varies, shown per row)**

| **Item** | **Summary** |
| --- | --- |
| Current role (n=29) | Histopathologist 31.03%; Dietitian 17.24%; Gastroenterologist 13.79%; GP 3.45%; Other 34.48% (incl. 1 oncologist) |
| Duodenal biopsies on same block/slide as other GI sites? (n=13) | No 92.3% (12); Yes 7.7% (1) |
| Proportion of endoscopic cases containing duodenum only (n=9) | 0%: 2; 10%: 3; 20%: 2; 40%: 1; 100%: 1 |
| Feasible to add 'duodenal endoscopy looks normal' flag? (n=10) | Yes for both endoscopy report & pathology form: 8; Yes, endoscopy report only: 2 |
| Importance of detailed microscopic description vs. diagnosis alone (n=19) | Very important 42% (8); Extremely important 21% (4); Slightly important 21% (4); Neutral 16% (3) |
| Would minimal AI-generated statement suffice for a report? (n=19) | No 52.6% (10); Yes 26.3% (5); Don't know 21.1% (4) |
| Typical pathologist time per duodenal biopsy (n=13) | 2–5 min 38.46%; 5–8 min 23.08%; <2 min 15.38%; >15 min 15.38%; 10–15 min 7.69% |
| Does centre scan all duodenal biopsies? (n=18) | Don't know 50% (9); No 38.9% (7); Yes 11.1% (2) |
| Minimum time saving needed for AI assistant to be worth introducing (n=10) | 2–5 min 40%; Less than 2 min 20%; 8–10 min 20%; 5–8 min 10%; 10–15 min 10% |
| Preferred scenario: A) full automation vs. B) assistant only (n=16) | B (assistant) 81.3% (13); A (full automation) 18.8% (3) |
| How content with AI reporting normal / coeliac / both automatically (n=16) | Distribution across Very content – Definitely not content; majority content for normal-only automation, least content for both combined |
| Usefulness of IEL/enterocyte count & VH:CD ratio - assistant model (n=15) | Majority rated Useful/Very useful for both metrics |
| Usefulness of same metrics - fully automated model (n=15) | Majority rated Useful/Very useful for both metrics, similar pattern to assistant model |
| Benefits of AI for normal-biopsy reporting (n=14, rank counts) | Reducing turnaround time, saving staff time and improving objectivity ranked most highly; saving money and future investment ranked lower; 1 free text 'other' (removing human responsibility) |
| Obstacles to AI for normal-biopsy reporting (n=14, rank counts) | Concerns about effectiveness and lack of digital infrastructure ranked highest; resistance from pathologists, referring clinicians and patients also cited; lack of guidelines and staff resources also raised |
| Consent to further contact (n=13) | Yes 38%; No (do not wish to be contacted further) 62% |

#### Supplementary Table 11

**Healthcare professional survey - free text comments**

**Explanation of preference for full automation (Scenario A):**

• If choosing the ‘pathologist's assistant’ option, it isn't giving a definitive answer - you would want a diagnosis if using AI.

• Remove human responsibility. Good luck.

• Cuts my time to evaluate and increased my diagnostic accuracy.

**Explanation of preference for assistant-only (Scenario B):**

• The tech could miss some other diagnosis.

• To begin with, AI-assisted biopsy reports should be signed out by a pathologist. Once the machine has been tested over a period of considerable time, the report may be generated by AI.

**Other benefits (free text):**

• Removing human responsibility.

**Other role (free text):**

• Oncologist.

#### Supplementary Table 12

**Healthcare professional workshop – demographics and outcomes**

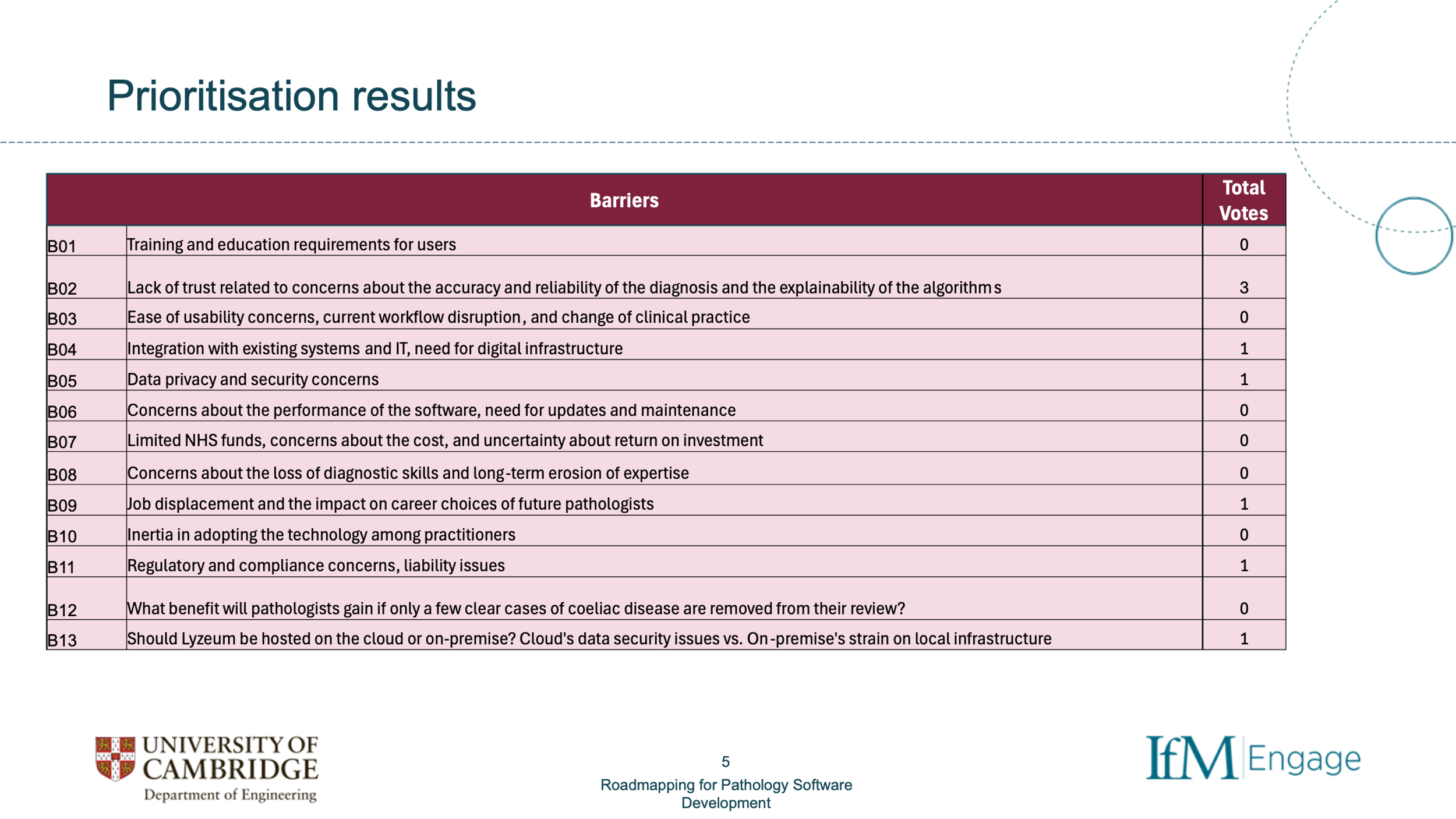
**Attendance: Seven healthcare professionals - one gastroenterologist, four gastrointestinal/general histopathologists (one of whom also held an NHS management role), and two further NHS managers. The session was facilitated by IfM Engage, a knowledge transfer company within the Institute for Manufacturing (IfM), which is part of the Department of Engineering at the University of Cambridge.**

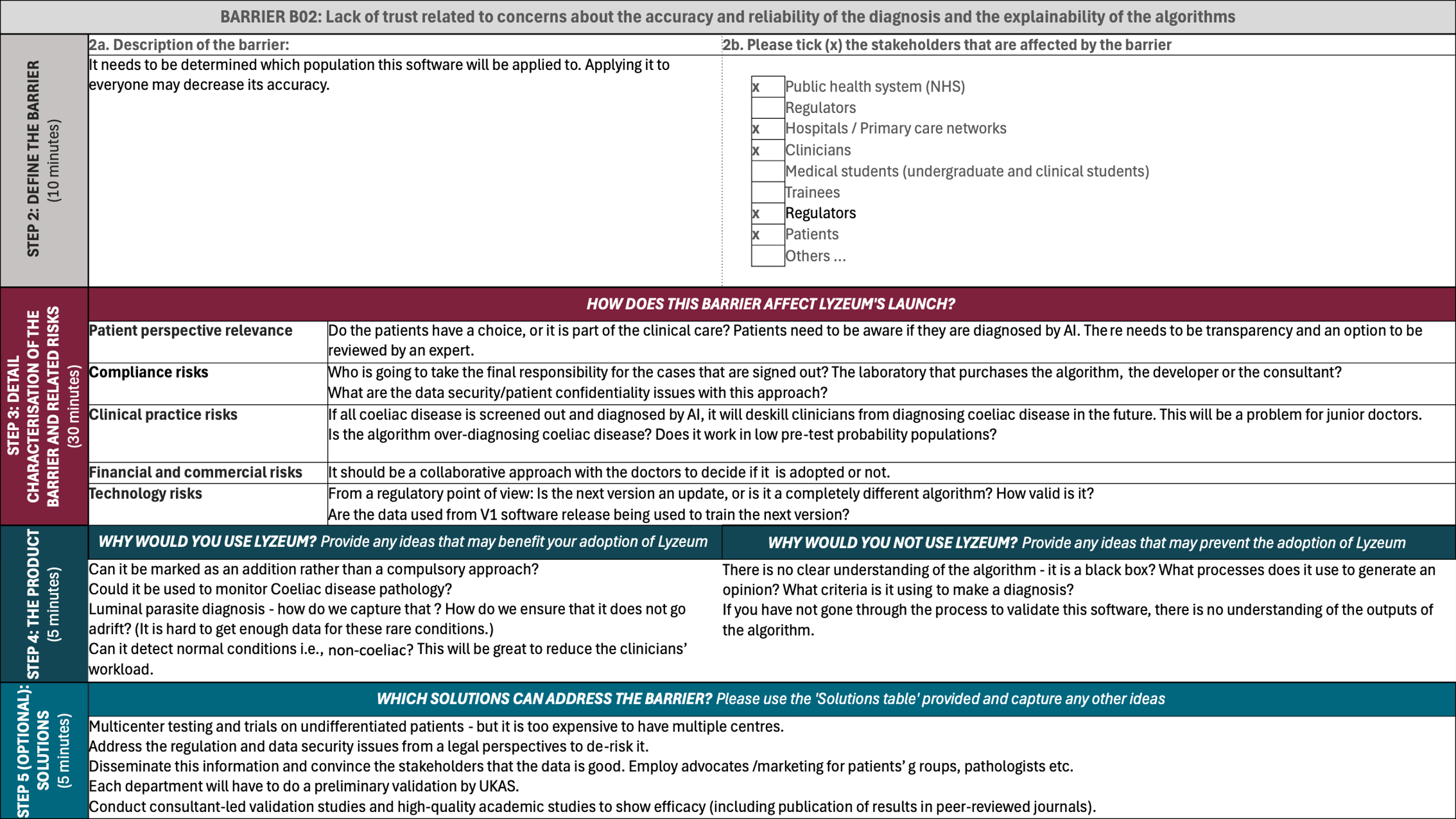

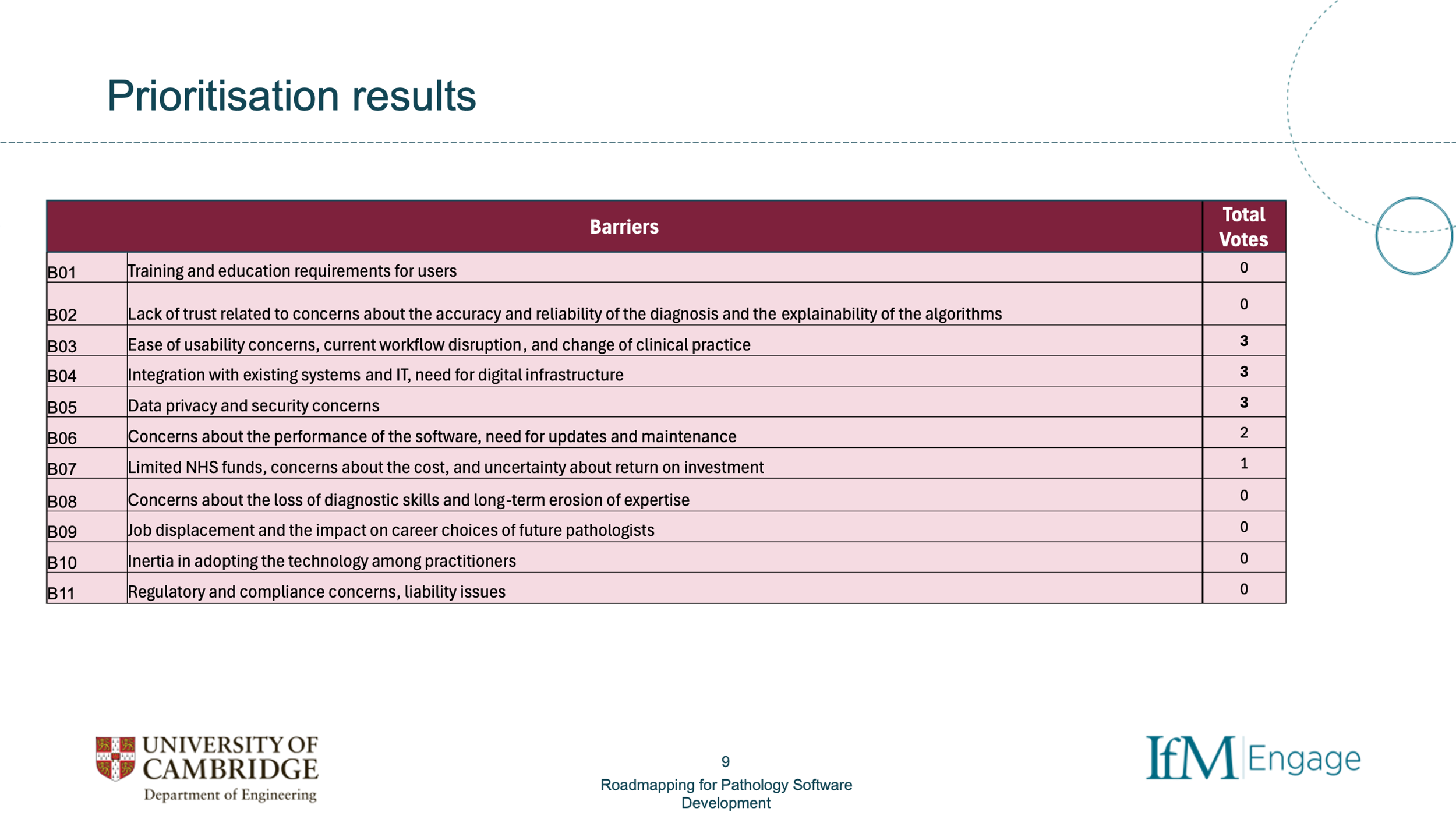

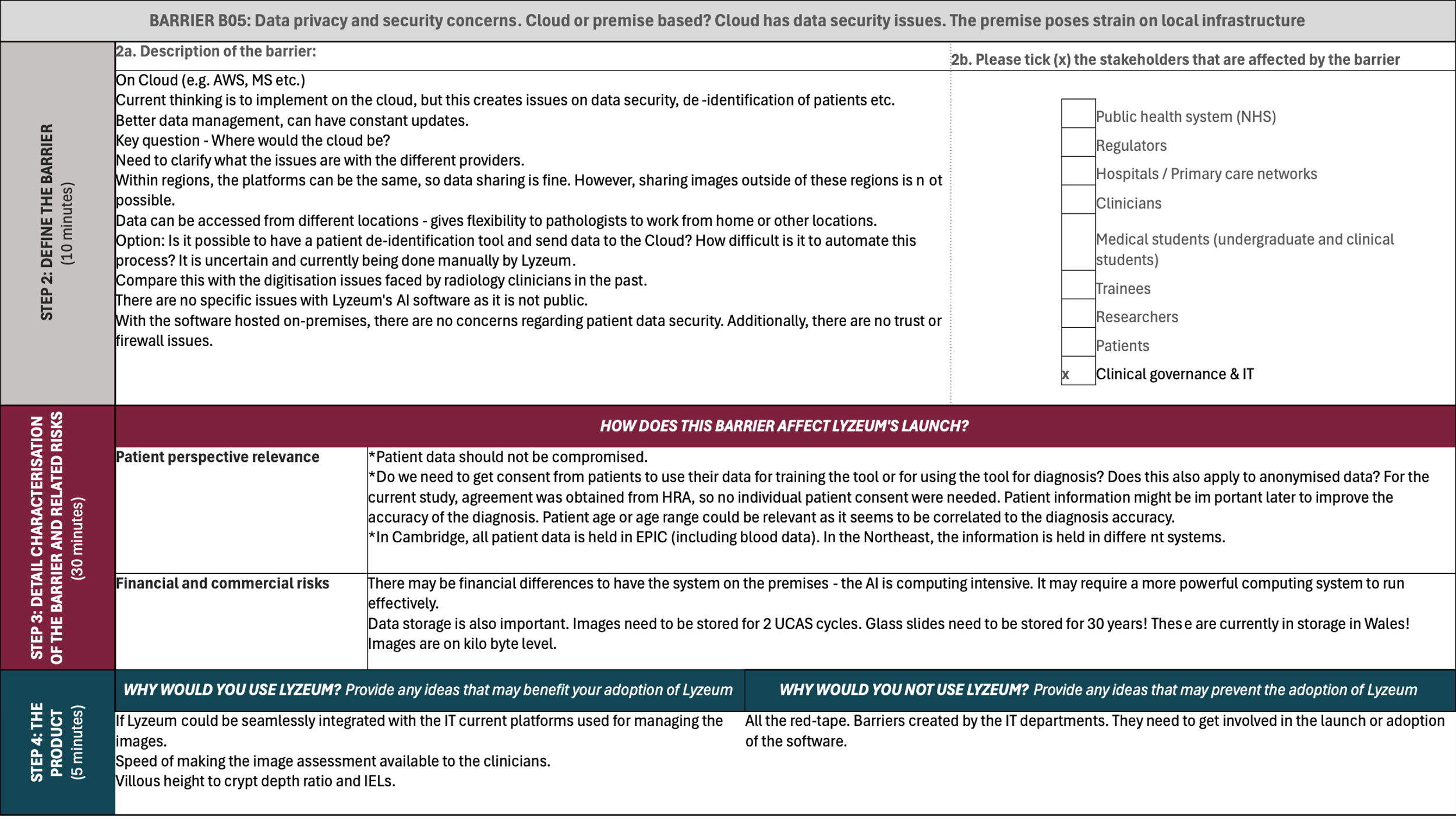

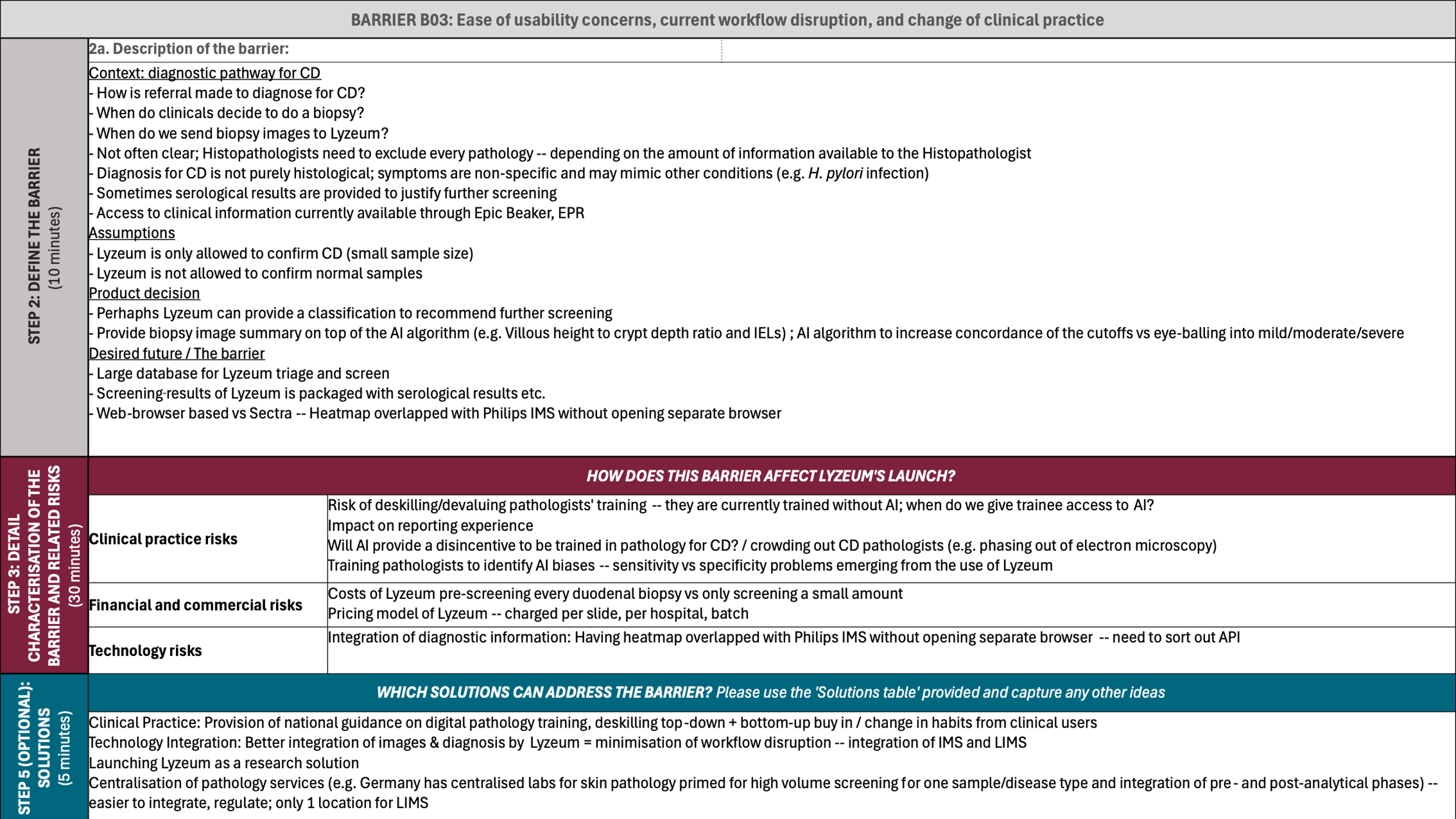

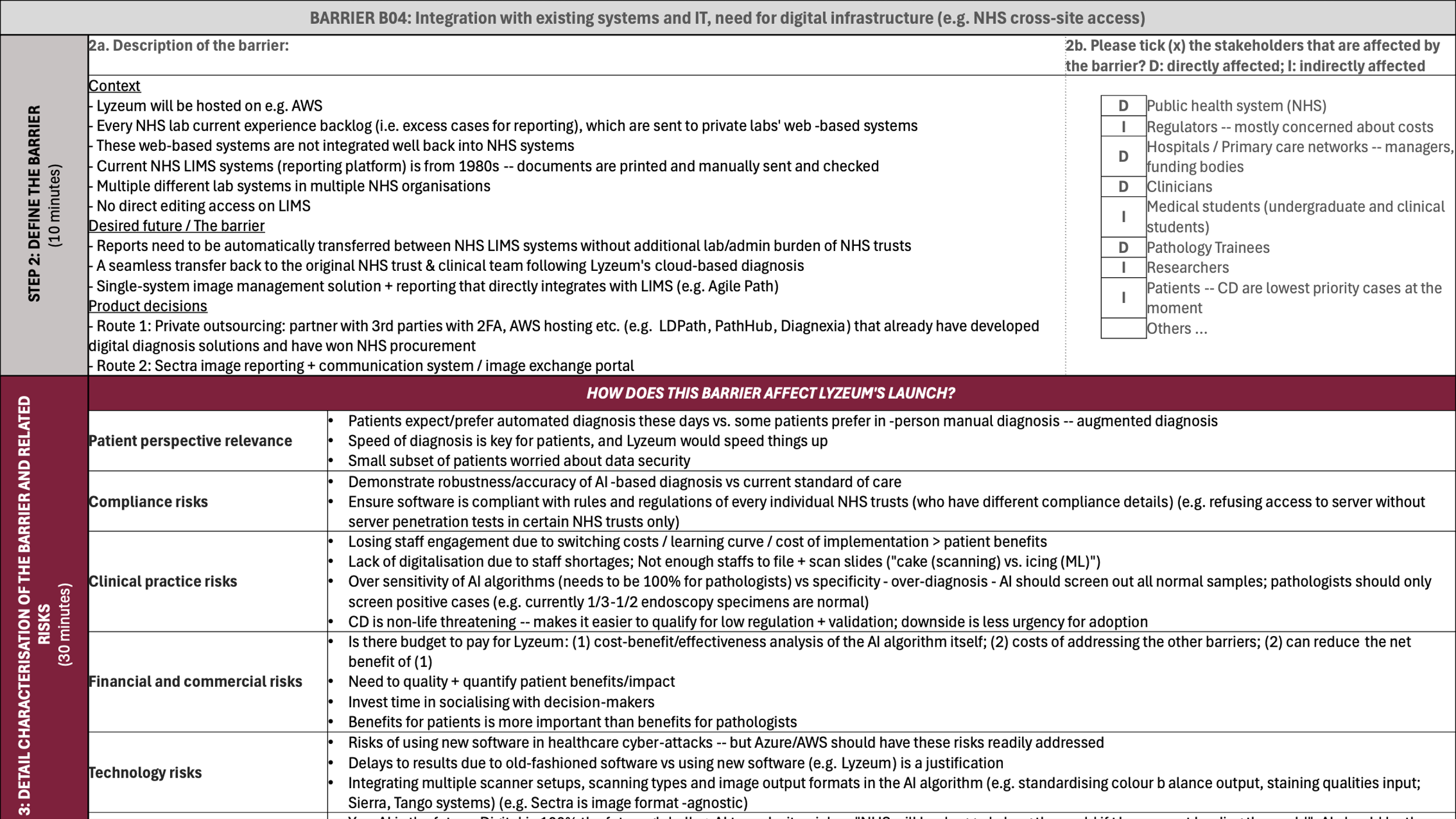
